# XBP1 expression in pancreatic islet cells is associated with poor glycaemic control across ancestries especially in young non-obese onset diabetes

**DOI:** 10.1101/2023.05.04.23289501

**Authors:** Theo Dupuis, Ranjit Mohan Anjana, Sundararajan Srinivasan, Adem Y Dawed, Alaa Melhem, Margherita Bigossi, Alasdair Taylor, Ebenezer Tolu Adedire, Jebarani Saravanan, Ambra Sartori, David Davtian, Venkatesan Radha, Sam Hodgson, Alison McNeilly, James Cantley, Naveed Sattar, Rohini Mathur, Sarah Finer, Genes & Health Research Team, Ewan R Pearson, Ana Viñuela, Rajendra Pradeepa, Viswanathan Mohan, Colin N A Palmer, Andrew A Brown, Moneeza K Siddiqui

## Abstract

**Objective:** Certain ethnicities such as South Asians and East Asians have higher rates of type 2 diabetes mellitus, in part, driven by insulin deficiency. Insulin deficiency can be due to beta-cell insufficiency, low beta-cell mass, or early cell death. Transcription factor *XBP1* maintains beta-cell function and prevents early cell death by mitigating cellular endoplasmic reticulum stress. We examine the role of *XBP1* expression in maintaining glucose homeostasis, glycaemic control, and response to diabetes therapeutics.

**Research Design and Methods:** Colocalisation analyses were used to determine if expression of *XBP1* in pancreatic islets and type 2 diabetes shared common causal genetic variants. We identify a lead eQTL variant associated exclusively with XBP1 expression and examine its association HOMA-B and stimulated glucose in cohorts of newly diagnosed Asian Indians from Dr. Mohan’s Diabetes Specialities Centre, India (DMDSC) and the Telemedicine Project for Screening diabetes and complications in rural Tamil Nadu (TREND). We then examine longer term glycaemic control using HbA1c in Asian Indian cohorts, the Tayside Diabetes Study (TDS) of white European ancestry in Scoltand, and the Genes & Health (G&H) study of British South Asian Bangladeshi and Pakistani ancestry. Finally, we assess the effect of eQTL variant on drugs designed to improve insulin secretion (sulphonylureas and GLP1-RA).

**Results:** Variants affecting *XBP1* expression in the pancreatic islets colocalised with variants associated with T2DM risk in East Asians but not in white Europeans. Lower expression of *XBP1* was associated with higher risk of T2DM. rs7287124 was the lead eQTL variant and had a higher risk allele frequency in East (65%) and South Asians (50%) compared to white Europeans (25%). In 470 South Asian Indians, the variant was associated with lower beta-cell function and higher stimulated glucose (β_log HOMAB_ =-0.14, P=5×10^-3^). Trans-ancestry meta-analysed effect of the variant in 179,668 individuals was 4.32 mmol/mol (95%CI:2.60,6.04, P=8×10^-7^) per allele. In 477 individuals with young onset diabetes with non-obese BMI, the per allele effect was 6.41 mmol/mol (95%CI:3.04, 9.79, P =2×10^-4^). Variant carriers showed impaired response to sulphonylureas.

**Conclusion:** *XBP1* expression is a novel target for T2DM with particular value for individuals of under-researched ancestries who have greater risk of young, non-obese onset diabetes. The effect of *XBP1* eQTL variant was found to be comparable with or greater that the effect of novel glucose-lowering therapies.

**Visual abstract:** Visual abstract: ER: Endoplasmic Reticulum, UPR: Unfolded Protein Response, IRE1:Inositol-Requiring Enzyme 1, mRNA: messenger ribonucleic acid, ERAD: Endoplasmic Reticulum Associated protein Degradation, eQTL: expression Quantitative Trait Loci, HbA1c: glycated haemoglobin. Created with Biorender.com

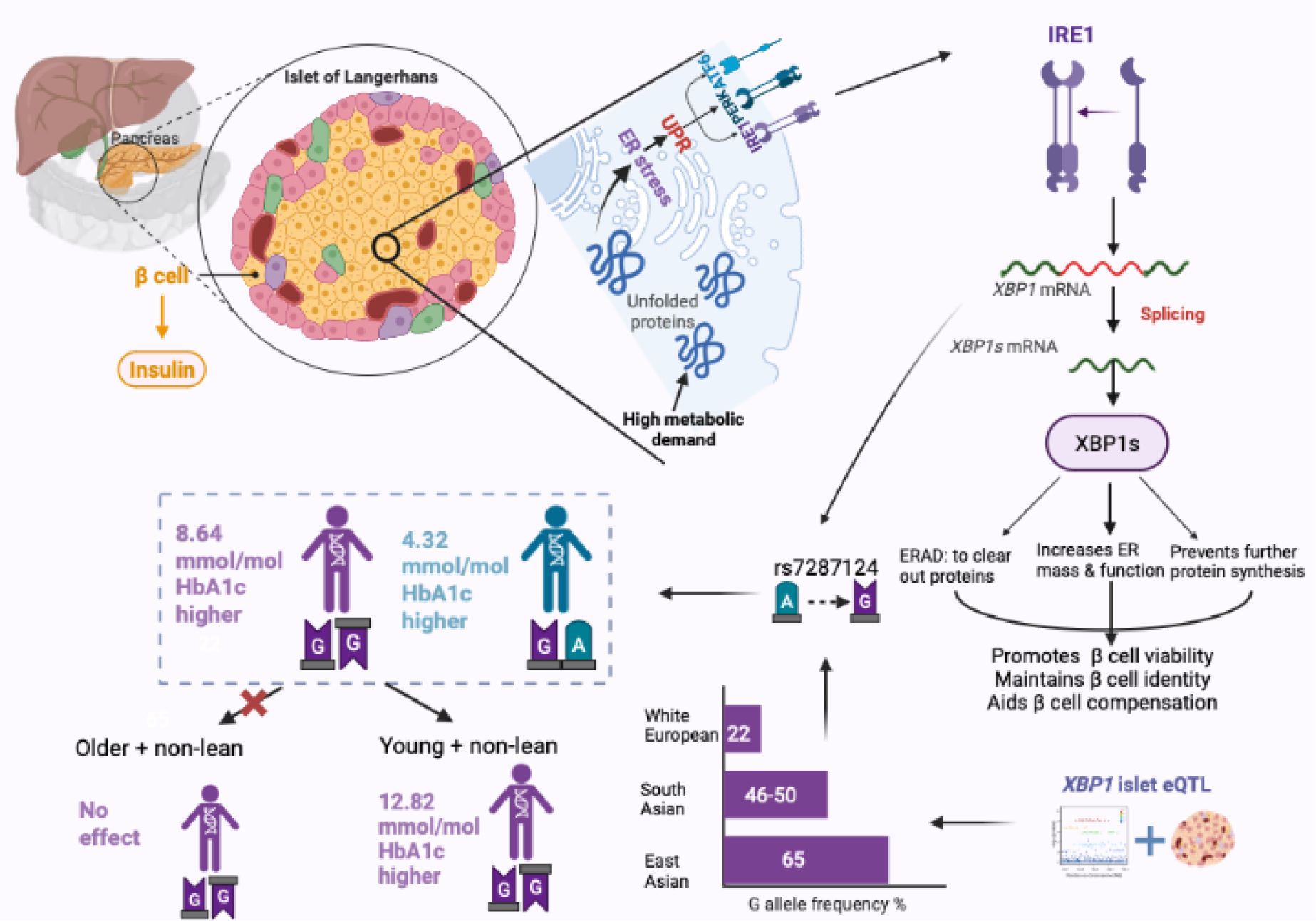

## Introduction

The pathophysiology of type 2 diabetes mellitus (T2DM) is multifactorial ^1^. The presentation of clinical features at diagnosis varies from those predominantly related to insulin resistance to insulin deficiency and mild age-related diabetes ^1,2^. While white Europeans show higher proportion of insulin resistance-related diabetes, those of South Asian, East Asian and African ancestry have a higher proportion of insulin deficiency ^2–6^. This, in part, leads to the higher frequency of young-onset diabetes in people of Asian and African ancestry ^7^. Insulin deficiency is driven by pancreatic beta-cell dysfunction, which could be due an insufficient number of beta cells, reduced mass, function, early cell death, increasing visceral fat or perhaps a combination of these factors.

The genetics of beta-cell function in humans is not as well studied as other aspects of T2DM. This is largely because most genetic studies of diabetes have been performed in well-powered cohorts of European ancestry, who have a lower burden of beta-cell dysfunction compared to other ancestries ^8^. Furthermore, the largest meta-genome-wide association studies (GWAS) on beta-cell function was performed in healthy white Europeans ^9^ and remains the only contributor even in recent studies ^10,11^. To address this gap in both data and knowledge, it is imperative to build on biological evidence of beta-cell susceptibility pathways and test their role in T2DM in non-white populations using approaches other than GWAS.

The efficiency of pancreatic beta-cells depends on a highly competent endoplasmic reticulum (ER) due to the high amount of protein synthesis and secretion required, especially in high-demand states ^12^. The unfolded protein response (UPR) is a key cellular process reversing ER stress ^13,14^. Therefore, an unimpaired UPR is crucial for continued insulin synthesis throughout life. Persistent ER stress, caused by impairments in UPR, has been linked to beta-cell dysfunction and death, while a functional UPR promotes increased insulin secretion ^13,15–18^.

XBP1 is a transcription factor produced in response to ER stress and its function is to regulate the UPR ^19,20^. Impairment of the UPR is linked to a decline in islet function in humans and mice; islets of eight individuals with T2DM showed decreased spliced *XBP1* expression ^21^. We elucidate this mechanism in **Figure 1**. Recently, Lee *et al.* showed that knocking out XBP1 in high-fat diet-fed mice resulted in the development of T2DM ^22^. They observed that these mice suffered from failed beta cell compensation and increased apoptosis. Pancreatic beta-cells in XBP1 knock-out mice showed greater beta-cell dedifferentiation, beta-to-alpha cell transdifferentiation, and impaired proinsulin processing. Together this rendered the beta-cells ineffective when challenged with metabolic stressors, leading to reduced beta-cell function and eventually hyperglycaemia. Summary statistics from the most recent trans-ancestry genome-wide association study of T2DM reported a credible set of 126 variants in the genomic region on chromosome 22, but so far none have been implicated with *XBP1* expression ^11^. The role of XBP1 expression in maintaining glucose homeostasis, glycaemic control, and response to diabetes therapeutics across human populations has not been previously studied.

**Figure 1.**
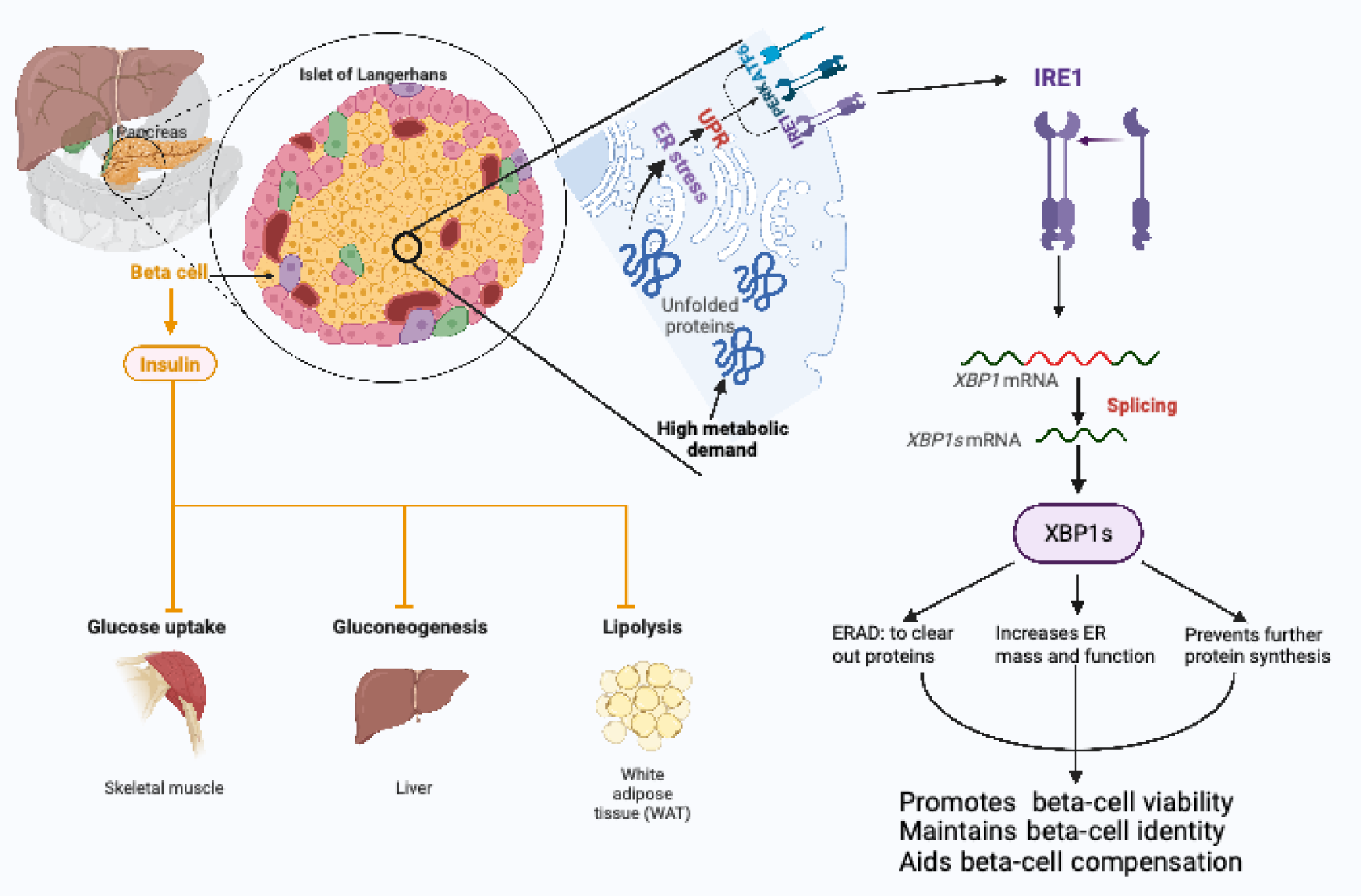
Role of XBP1 in regulating endoplasmic reticulum stress in pancreatic beta cells. Created with BioRender.com

We test the hypothesis that an eQTL (variants that alter the expression of a gene) for *XBP1* in pancreatic islet cells are associated with increased risk of T2DM, reduced beta-cell function (HOMA-B), and impaired pre-treatment glycaemic control (HbA1c). We also assess whether this genetic region has been under population selection in Europeans who show lower prevalence of beta-cell dysfunction.

## Methods

### Identifying potentially causal variant for pancreatic islet expression of XBP1

To identify lead eQTL variant, we used expression data from pancreatic islets (n=420) and confirmed its direction of effect in pancreatic beta cell samples (n=40) from the Integrated Network for Systematic analysis of Pancreatic Islet RNA Expression (InsPIRE) study (details in **Supplementary Methods 1**). To rule out false positive or pleiotropic effects, we examined the variant in the Translational human pancreatic Islet Genotype tissue-Expression Resource (TIGER) ^23^. A study flowchart is provided in **Supplementary Figure 1**.

### Colocalisation analyses for XBP1 expression and T2DM risk

We performed colocalisation analyses to ensure that the lead variant identified was associated with both expression of *XBP1* in pancreatic islets and type 2 diabetes risk. Colocalisation analyses compare the distribution of summary statistics from large-scale association studies of complex traits and eQTLs, while reducing false positive discoveries by correcting for linkage disequilibrium across a genetic region ^24,25^. For colocalisation analyses, we used the programme, ezQTL, which computes the probability of colocalisation using two methods, HyPrColoc (Hypothesis Prioritisation in multi-trait Colocalization) ^26,27^ and eCAVIAR (CAusal Variants Identification in Associated Regions) ^28^. Since, colocalisation analyses was used to check for linkage contamination, only single-ancestry GWA studies could be utilised. Therefore, we used summary statistics from the two largest available ancestry-specific GWAS of T2DM: one of white Europeans and the other of East Asian descent from the DIAMANTE consortium and Biobank Japan respectively. We then assessed the direction of effect between expression and T2DM risk using inverse variance weighted and weighted median methods in the MR-Egger package ^29^.

All publicly available resources used for summary statistics have been published on previously, additional details on these are provided in **Supplementary Table1, Methods1.**

### Association of XBP1 expression with glycaemic traits

We used individual-level data from 4 biobanks in this analysis: the Tayside Diabetes Cohort based in Scotland, UK (TDC, comprised of white Europeans), Dr. Mohan’s Diabetes Specialties Clinic and Telemedicine Project for Screening diabetes and complications in rural Tamilnadu (DMDSC & TREND, both comprised of South Asian Indians based in Tamilnadu, India), and finally Genes & Health (G&H, British South Asian Pakistani and Bangladeshi) based in London, UK. Study design and data collection for these biobanks have been described previously and additional details in **Supplementary Methods2.** ^3,30–33^. Briefly, TDC and G&H are cohorts with record linkage of electronic health data from their local National Health Service boards which have been used for ancestry-specific diabetes research ^3,34^. DMDSC have record linked clinical data from visits to a private diabetes clinic and hospital chain in south India. In TDC, G&H and DMDSC only participants with newly diagnosed T2D (up to 12 months before and 1 month after data collection) were included. TREND is a survey of diabetes prevalence in rural south India. In TREND, only participants with screen-detected diabetes only were considered. Additional details about the cohorts including genotyping methods are provided in Supplementary Methods. All studies used WHO criteria for diagnosis of T2DM using fasting or random plasma glucose, OGTT, or glycated haemoglobin (Hba1c) ^35^. Serum C peptides (where available), fasted and stimulated glucose, and HbA1c were restricted to measures taken 12 months prior to and up to 1 month after diagnosis of T2DM to reduce contamination of effects by diabetes therapies.

### Glycaemic traits

Serum C-peptides and plasma glucose were measured only in the DMDSC biobank and using a fixed protocol fasted and stimulated measures ^36^. Patients visit the facility in a fasted state and a blood draw is made. These are then used in Homeostatic Models of Assessment (HOMA) to estimate pancreatic beta-cell function and insulin sensitivity ^37,38^. Similarly, we assessed the effect of the eQTL variant on stimulated blood glucose (2hrG) levels in rural-dwelling Asian Indians who were screen-detected as having diabetes during the TREND survey.

HbA1c is commonly recorded and provides a measure of long-term blood sugar control. We tested the association of the variant rs7287124 with HbA1c from all four biobanks. For meta-analyses of the effect of rs7187124 with HbA1c in people without diabetes, we utilised summary estimates from the Meta-Analysis of Glucose and Insulin-related traits Consortium (MAGIC). Summary estimates from the three largest ancestry-specific contributors: Europeans(70%), East Asians(13%), and South Asians were used ^10^.

### Sub-group analyses

Lower *XBP1* expression impacts beta-cell viability under high metabolic stress. Therefore, we hypothesized that the effects of the variant would vary by BMI and age at diagnosis, and this would be detectable in an interaction term between. We include an interaction variable to control for confounding effect in full models. Additionally, sub-groups were defined guided by the interaction effects.

Thresholds for non-obese and obese BMI followed ethnicity-specific guidelines; non-obese for white Europeans was BMI <30 kg/m^2^, whereas for South Asian Indians <25 kg/m^2^ and South Asian Pakistanis and Bangladeshis living in the UK < 27.5 kg/m ^39–41^. Associations between the *XBP1* eQTL variant were tested in the full cohort, and in those with 1) young onset and non-obese BMI at diagnosis and 2) in those with older onset and obese BMI. Genetic data from these cohorts has been used previously, an overview of genotyping methods is provided in **Supplementary Methods 3.**

### Statistical models used

For HOMA-B associations we used the following 3 models: one unadjusted model assessing genotype effect on beta-cell function, second adjusted for HOMA-S (insulin sensitivity), age, BMI, and sex, and a third to determine if there was an interaction between the variant age and BMI at diagnosis. For stimulated blood glucose models were adjusted for BMI and age at diagnosis as well as sex. Finally, for HbA1c association testing, the full model was:

HbA1c ∼ eQTL variant + Age at diagnosis + BMI at diagnosis + Sex + eQTL variant*Age at diagnosis* BMI at diagnosis

Sub-group analyses models were not adjusted for the interaction, the model below was used: HbA1c ∼ eQTL variant + Age at diagnosis + BMI at diagnosis + Sex

All analyses were undertaken using R4.2.1^42^. We meta-analysed the overall and sub-group stratified associations assuming random-effects using the R package Metafor ^43^

### Pharmacogenomic effects on sulphonylurea and GLP1-RA response

Finally, to test the hypothesis that XBP1 expression exerts and effect on glycaemic control through beta-cell function, we used the largest available pharmacogenomic studies from the MetGen Plus and the DIRECT consortia to examine the effect of the eQTL variant on response to an incretin mimetic like glucagon-like peptide 1-receptor agonist (GLP-1RA) and insulin secretagogues like sulphonylureas ^44,45^.

## Results

### Rs7287124 is the lead eQTL variant for XBP1 expression in pancreatic islets

We identified a single intronic variant; rs7287124 (Chromosome 22: 29239157A>G) is a *cis*-eQTL for *XBP1* expression in pancreatic islets (β _expression_ = −0.18, p = 2×10^-4^ (Table 1). The direction of expression effect was consistent in pancreatic beta cells (N = 26, β _expression_ = - 0.219, p = 0.54). Using TIGER summary statistics, we confirm that the G allele is associated with lower *XBP1* expression **(Table1 & Supplementary Table 2**) and is not an eQTL for any other genes. Further, the effect of rs7287124 in both Biobank Japan ^46^ and the DIAMANTE trans-ancestry meta-GWAS summary estimates ^11^, is directionally consistent with increased diabetes risk **(Table 2)**. The EAF in European populations is 24%, whereas South Asians and East Asians have higher EAF of 54% and 65% respectively, suggesting that these ethnicities will have lower genetically predicted *XBP1* levels in pancreatic islets.

**Table 1.**
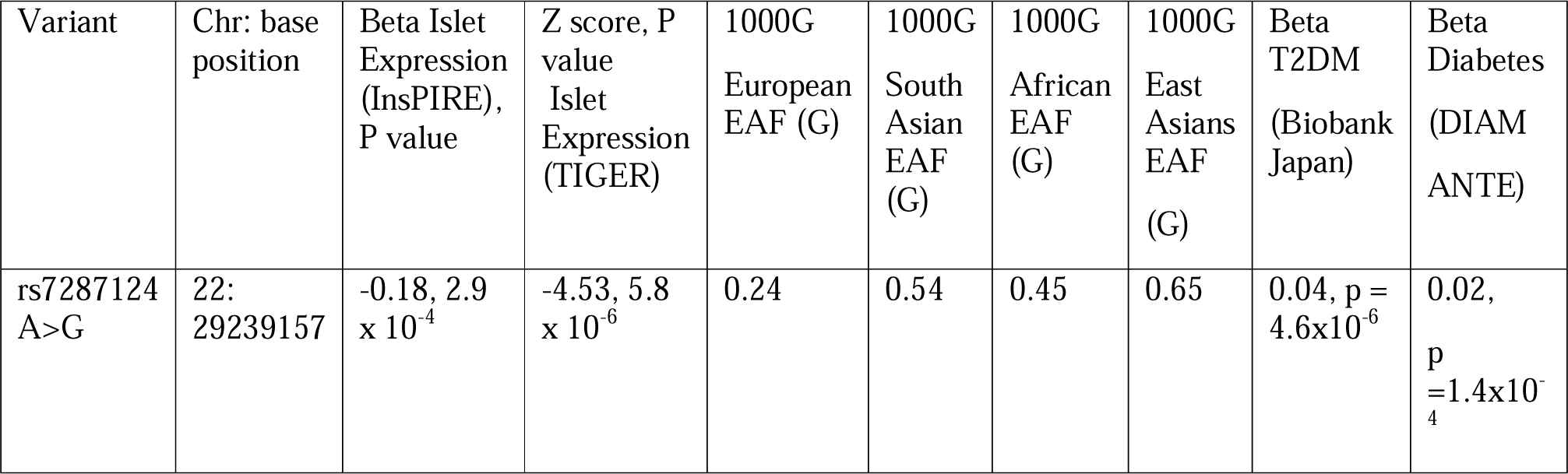
Allelefrequencies across ancestries and effects of lead eQTL variant on XBP1 expression in pancreatic beta-cells, T2DM risk.

| Variant | Chr: base position | Beta Islet Expression (InsPIRE), P value | Z score, P value Islet Expression (TIGER) | 1000G European EAF (G) | 1000G South Asian EAF (G) | 1000G African EAF (G) | 1000G East Asians EAF (G) | Beta T2DM (Biobank Japan) | Beta Diabetes (DIAM ANTE) |
| --- | --- | --- | --- | --- | --- | --- | --- | --- | --- |
| rs7287124 A>G | 22: 29239157 | -0.18, 2.9 x 10<sup>-4</sup> | -4.53, 5.8 x 10<sup>-6</sup> | 0.24 | 0.54 | 0.45 | 0.65 | 0.04, p = 4.6x10<sup>-6</sup> | 0.02, p = 1.4x10<sup>-4</sup> |

### Colocalisation analysis suggests shared causal variant between *XBP1* expression and T2DM risk

In order to rule out linkage contamination, i.e. the possibility that variants other than rs7287124 were driving the effects on T2D risk, we undertook colocalisation analyses using the ezQTL platform. We found a strong posterior probability using HyPrColoc and eCAVIAR, that the same variants are associated with T2DM risk and *XBP1* expression in East Asians (**Figure2a**), while weaker probabilities were observed in white Europeans. Summary statistics are available in **Supplementary Figures2-4**). Colocalisation analyses strongly suggest that *XBP1* expression, rather than that of any of the surrounding genes, is causally linked with T2DM risk. Both methods identified variants in the gene *ZNRF3*, which neighbours *XBP1* and contains eQTLs for *XBP1*.

We validate the *ZNRF3*’s involvement in T2DM by examining evidence for genetic burden of common variants located in the gene using the <u>AMP-T2DM knowledge portal</u>. Using summary statistics of all GWAS available on the knowledge portal, the Human Genetic Evidence guidelines (HuGE) tool calculates a Bayes Factor of 45 x 5^-8^ concluding that there is very strong evidence for the involvement of variants located in this gene with T2DM risk in humans (**Supplementary Figure5**) ^47^. Interestingly, a test for the well-known beta-cell gene *TCF7L2* produces a similar level of evidence.

The variant identified in the colocalisation analyses (rs58004020) is in strong LD with the eQTL variant rs7287124 in South Asians, East Asians and Europeans: R^2^=0.98, 0.99 and 0.98 respectively while D’=0.85, 1 and 0.72 respectively. Both variants are in *ZNRF3*. We then investigate the direction of effect by testing the hypothesis that lower *XBP1* expression in pancreatic islets was associated with higher T2DM risk. Using inverse variance weighted and weighted median models, we find this association between the two traits to be significant (B −0.065, SE 0.02 and P< 7.17×10^-4^) and directionally consistent with our hypothesis (**Figure 2b**).

**Figure 2.**
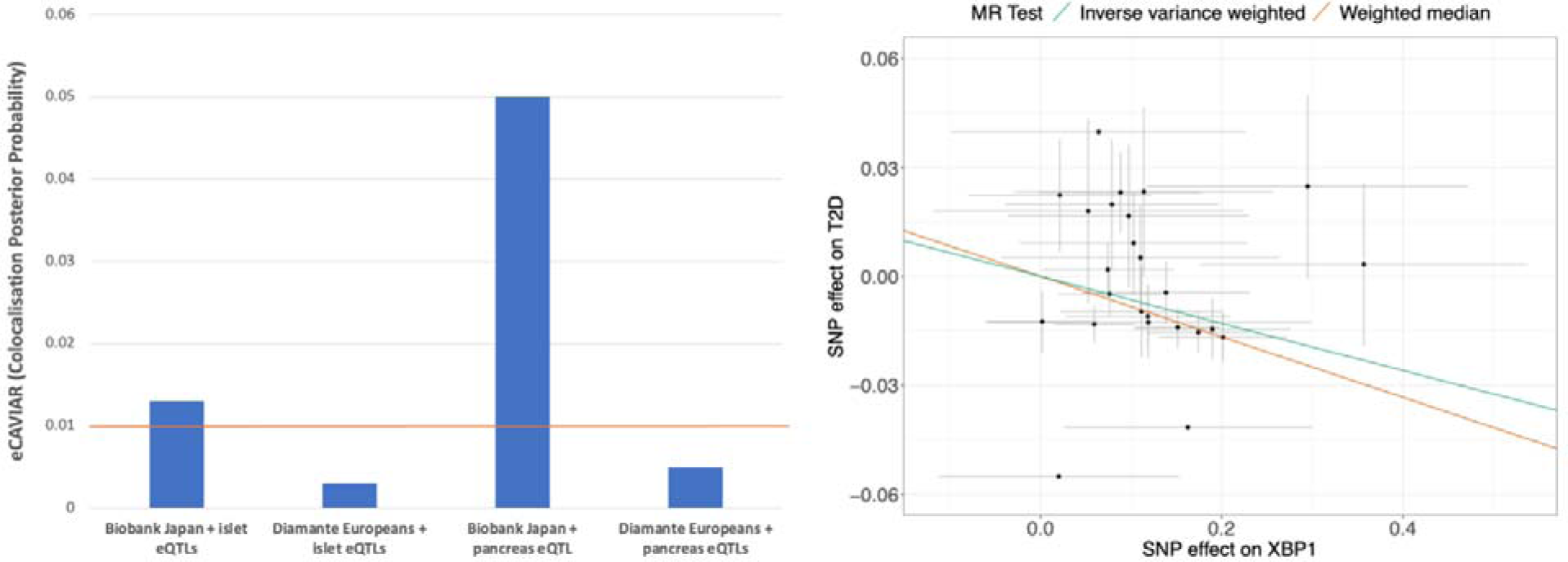
a) Colocalisation probabilities using eCAVIAR and HyPrColoc between XBP1 expression in both pancreatic islets and GTEx pancreas samples and type 2 diabetes risk in Biobank Japan and white Europeans from DIAMANTE. Orange line indicate eCAVIAR recommended threshold for significant colocalisation. In both cases, East Asians are more likely to have overlapping causal variants. b) Ratio estimates of association between XBP1 expression in pancreatic islets and T2D risk in Biobank Japan. These plots have been made using 25 variants with LD < 0.4 in the XBP1 region +/-5KB. Weighted median estimate for the effect was −0.083 (SE: 0.025), P value =9.2 x 10^-3^, and inverse variance weighted estimate was −0.065 (SE: 0.020), P value = 7.17 x 10 ^-4^.

Therefore, rs7287124 was selected for downstream investigation with glycaemic traits and drug response.

### Association with human beta-cell function in Asian Indians

Baseline characteristics of cohorts with individual-level data are described in **Supplementary Table4**. In 470 participants of the Asian Indian cohort, the *XBP1* eQTL variant rs7287124 was associated with HOMA-B in unadjusted (**Figure3a**) and adjusted models (**Supplementary Table5**). In additive models of genetic effect adjusted for HOMA-S (insulin sensitivity), age, BMI and sex, the G allele was associated with lower HOMA-B (B: −0.14, SE: 0.05, P=5×10^-3^). An interaction between the rs7287124, age, and BMI at diagnosis was observed (P=4.3 x 10^-5^). In individuals diagnosed young and non-obese, this effect was greater (n=82, B: −0.30, SE: 0.14, P = 0.036). However, the effect was not significant in the older and non-lean sub-group (n=134, P=0.27). Recessive models were run to confirm the genetic model is additive (**Supplementary Table5**). The variant was not associated with insulin resistance or sensitivity (P=0.12).

### Association with stimulated blood glucose levels

We then assess the association between rs7287124 and stimulated blood glucose (SBG) levels in 484 rural-dwelling Asian Indians from the TREND. While the variant was not associated with FBG, it was associated with higher SBG in unadjusted models (**Figure 3b**) (B=0.7 mmol/L, SE=0.3, P=0.03) in models adjusted for BMI, age, and sex. However, no effect was observable in either of the sub-groups.

**Figure 3.**
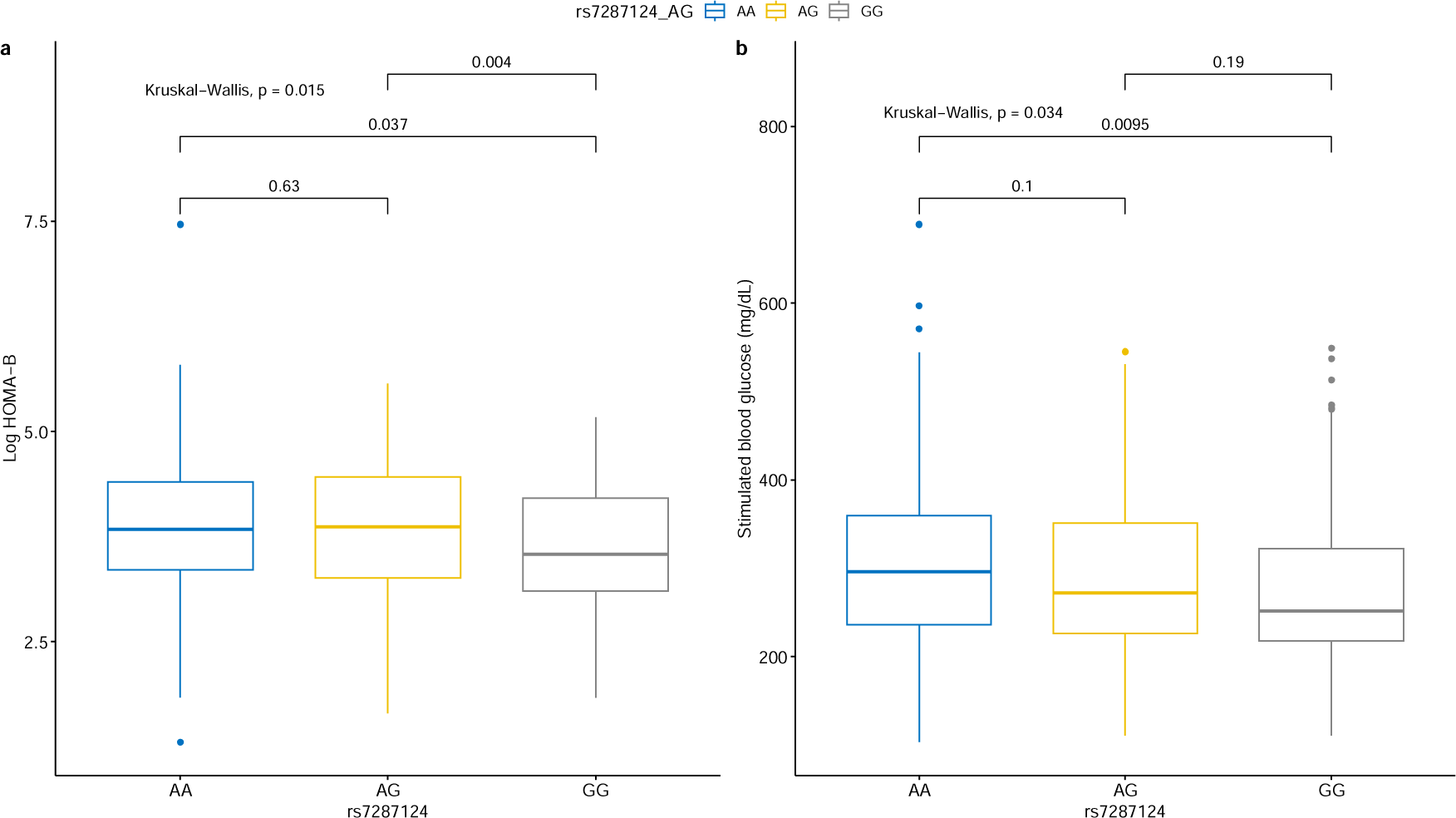
a) Box plot showing additive models of XBP1 eQTL variant (rs7287124) and HOMA-B calculated using fasted glucose and stimulated C-peptides in 470 individuals with newly diagnosed type 2 diabetes at DMDSC, Chennai India. After adjustment for age, sex, BMI and HOMA-S the association remained significant (P=5×10^-3^). b). Boxplot showing effect of rs7287124 on 2hr stimulated blood sugar in rural-dwelling Indians from the TREND study. After adjustment for age, sex and BMI the association remained significant (P=0.03). Per allele was associated with 0.7 mmol/L higher 2hr glucose.

We then tested the hypothesis that the variant will likely to be associated with longer term glycaemic control using HbA1c which is much more routinely measured.

### Association with HbA1c levels, especially in individuals diagnosed young and with non-obese BMI

To assess the effect of the XBP1 eQTL variant, rs7287124, across ancestries we included individual-level data from newly-diagnosed South Asian Indians represented by DMDSC (n=459) and TREND (n=471), South Asian Bangladeshi and Pakistani represented by Genes & Health (n=644) and, white Europeans from the TDC (4,908). Additionally, we used summary statistics from existing GWAS in individuals without T2DM.

The variant was in Hardy-Weinberg equilibrium in all cohorts. The trans-ancestry meta-analyses showed a significant increase in HbA1c per risk allele regardless of diabetes status **(Figure 4a).** Random-effects meta-analysis showed the pooled effect across T2DM status to be 4.32 mmol/mol per risk allele (95%CI: 2.60, 6.04), P value 8×10^-7^. Therefore, homozygous carriers of the G allele are estimated to have 8.64 mmol/mol higher HbA1c levels compared to non-carriers. In meta-analyses of the effect in healthy individuals, each risk allele was associated with 5.37 mmol/mol higher HbA1c (95%CI:0.22, 10.52), while in people with T2DM, the effect was 4.37 (95%CI:2.60, 6.55).

**Figure 4.**
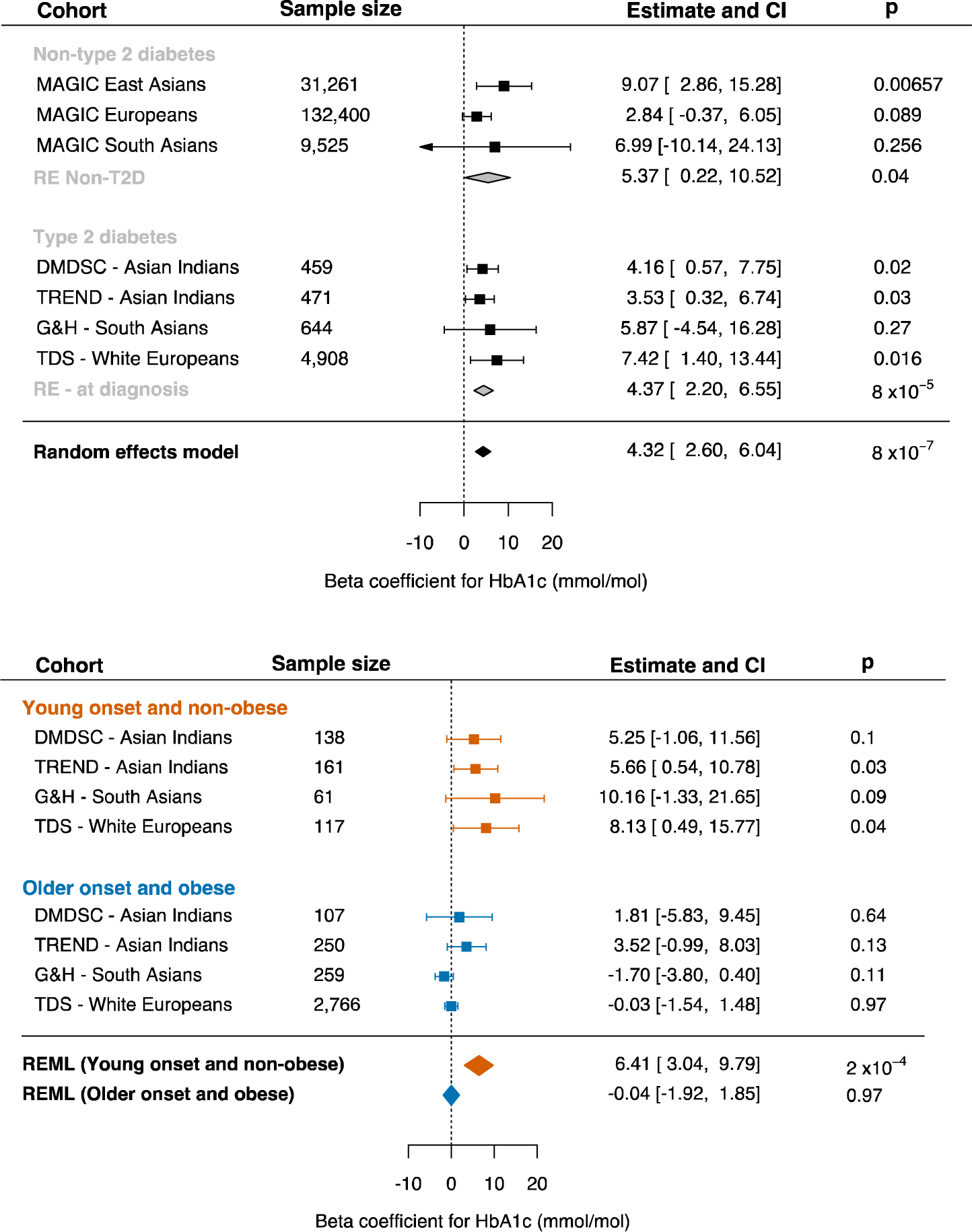
Forest plots showing random effects of XBP1 eQTL variant on HbA1c (mmol/mol) Effects are pooled across summary statistics available from the MAGIC consortium for East Asians, Europeans, and South Asians without T2DM (n=173,186) and for 6482 individuals with newly-diagnosed T2DM across 4 cohorts: Tayside Scotland (TDS), urban-dwellingSouth Asian Indians (DMDSC), a rural-dwelling South Asian Indians (TREND) and British South Asian Pakistani and Bangladeshis (G&H). Random effect meta-analysis shows an increase of 4.32 mmol/mol of HbA1c per risk allele (95CI: 2.60, 6.04). Forest plots of sub-groups of people with newly diagnosed T2DM showed a stronger effect in 477 individuals diagnosed young with non-obese BMI (6.41 mmol/mol), compared to no effect in those diagnosed older with non-lean BMI (n=3382).

Approximately 7% of individuals (n=477) with newly-diagnosed diabetes had young onset (<40 years) with non-obese BMI. Despite using ethnicity-specific BMI thresholds, this proportion differed across cohorts: in Asian Indians: 30% in DMDSC, 34% in TREND (<25kg/m2), in British South Asians from G&H 9.5% (<27.5 kg/m2), and in white Europeans from TDS 2.4% (<30 kg/m2). The meta-analysis in sub-groups shows that the effect of this variant is driven by the young, non-obese sub-group. In this group the per allele effect was 6.41 mmol/mol (3.04, 9.79), P = 2×10^-4^ **(Figure 4b).** In contrast, those diagnosed older with obese BMI (n=3,382) showed no genetic effect. In our study cohorts, 25% of South Asian Indians (DMDSC+TREND), 20% of South Asian Pakistani and Bangladeshi (G&H), and 0.05% of white Europeans (TDC) were homozygous carriers of the risk allele (G/G) (**Supplementary Figure6**).

### rs7287124 is associated with worse response to sulphonylureas but no differential response to GLP1-RA

Carriers of a variant allele associated with reduced beta-cell function are likely to have limited response to insulin secretagogues. To test this hypothesis, we examined the effect of these variants on glycaemic response to sulphonylureas and GLP-1RA. Carriers of the G variant showed no difference in response to GLP-1RA compared to non-carriers (**Supplementary Table6**). For sulphonylureas, we find that the variant allele was associated with a worse response, with each G allele associated with 0.061% (0.7 mmol/mol) lesser reduction in HbA1c (SE: 0.023, P value =0.008).

## Discussion

We find T2DM risk is associated with eQTLs exclusive for *XBP1* and that decreased expression of *XBP1* increases T2DM risk. We identify an eQTL for *XBP1* in pancreatic islets and beta-cells: rs728712(A>G). Rs7287124 is associated with lower beta-cell function using HOMA-B and higher stimulated glucose in Asian Indians with newly-diagnosed T2DM. In a trans-ancestry meta-analysis, we observed that variant carriers had higher HbA1c at diagnosis (P value 8×10^-7^), and that this effect was driven by individuals diagnosed young with non-obese BMI (P = 2×10^-4^). The variant is more common in those of South Asian (46-50%), East Asian (65-68%), and West African (45%) ancestry than in Europeans (22-25%). Consistent with risk allele frequency and previous epidemiological studies, we observed that the frequency of young and non-obese onset diabetes was higher in South Asian populations than in Europeans ^6,7^. Finally, we observed that treatment with insulin secretagogue: sulphonylurea was associated with higher-post treatment HbA1c in rs7287124 carriers.

### Clinical impact of genetic effect

To examine pre-treatment glycaemic control in individuals with new-onset diabetes we applied strict inclusion criteria around time of diagnosis. Our trans-ancestry meta-analysis showed the average per allele effect of rs7287124 on HbA1c was 4.32 mmol/mol (P=8×10^-7^). This is equivalent to an 8.64 mmol/mol (0.79%) difference when comparing homozygotes (those with AA v. GG genotypes). Individuals diagnosed young and with non-obese BMI the per allele effect was 6.41 mmol/mol which is equivalent to a difference of 12.82 mmol/mol (1.2%) in HbA1c comparing homozygotes.

All else being equal (age at diagnosis, sex, BMI and ethnicity), the per allele difference for rs7287124 was equivalent to or greater than newer diabetes therapies. A meta-analysis of the efficacy of recently introduced diabetes therapies showed the average reduction in HbA1c in response to DPP4-inhibitors, SGLT2is and GLP1-RA was 0.53%, 0.79%, and 0.78% respectively. These differences are more modest than the observed effect of rs7287124 on HbA1c in this study; particularly in the young and non-obese sub-group.

Use of candidate gene approach and importance of ancestrally diverse genetic studies The effect size observed in our study is not unusual, from the most recent trans-ancestry meta-GWAS of HbA1c, the largest effect size for common variants (such as rs2971669 in *GCK*) with similar allele frequency as rs7287124 (in Europeans), was ∼3.3 mmol/mol, which is similar to the effect observed in our study. However, signals such as *XBP1* are likely to be masked in such GWAS, as research in populations particularly susceptible to insulin deficiency or beta-cell dysfunction (such as South Asians, East Asians and west Africans) are limited. While the contribution of cohorts of non-European ancestries has been growing, studies of glycaemic traits are often still more strongly representative of Europeans (70%) ^8,10^. Similarly, pharmacogenomic studies that are mostly available from European cohorts. Finally, due to the challenge of capturing treatment-naïve glycaemic measures, large-scale meta-GWAS for glycaemic traits are undertaken in healthy individuals who do not have diabetes. The lack of individual-level data for these studies, particularly around the age and BMI at diagnosis, precludes them from sub-group analyses.

Therefore, this study utilises a candidate gene approach building on a strong foundation of *in vitro* and *in vivo* studies. To support this approach, we validate our target using colocalisation analyses with cell-specific transcriptomics, validating the signal in both available resources of pancreatic islet expression data in humans (InsPIRE and TIGER). In clinical studies across different ethnicities, we confirm the association of this variant with three glycaemic traits: HOMA-B, blood glucose and most robustly with HbA1c. We then validate the beta-cell depletion hypothesis by undertaking look-ups in pharmacogenetic studies of insulin secretagogues. Rs7287124 is also associated with lower haemoglobin and haematocrit levels HbA1c often under-represents the degree of dysglycemia in populations with lower hemoglobin ^48^.

Our approach is supported by colocalisation analyses which showed T2DM risk in East Asians rather than white Europeans was associated with *XBP1* expression. We observe the same pattern whether using eQTLs in the pancreas or islet cells. The stronger probability with pancreas samples is likely due to the larger single-study in GTEx relative to InsPIRE which had several contributing studies. The lower risk allele frequency in Europeans could be a result of positive selection. Indeed, calculating a gene-based score for recent selection based on data from the 1000 Genomes Selection Browser ^49^, we find *XBP1* (and a 20kb region flanking it which includes *ZNRF3*) is ranked in the top 6.8% of all genes and long non-coding RNAs in terms of evidence of recent selection in the European CEU population relative to the Chinese CHB population **(details in Supplementary 6).** Selection conversion suggests that protective alleles for increased *XBP1* expression have been selected for to a greater degree in Europeans, who also have a lower risk of beta-cell dysfunction and therefore lower frequency of young and lean onset diabetes. This would explain why risk of lower *XBP1* expression in colocalised with T2DM risk in East Asians rather than white Europeans.

### Pharmacogenetics validate hypothesis of beta-cell depletion

Sulphonylureas, which close the K_ATP_ channel and force beta-cells to secrete insulin independently of glucose concentrations, may place an additional load on the ER and secretory pathway. Moreover, sulphonylureas have been demonstrated to directly induce apoptosis in human islets^50^. We find that reduced islet *XBP1* levels in carriers of rs7287124 likely render beta-cells more sensitive to ER stress and therefore have diminished glycaemic control in response to sulphonylurea therapy. Future studies will be required to investigate the impact of sulphonylurea treatment on beta-cell function, differentiation, and survival in risk allele carriers.

GLP-1RA response is possibly limited by the fact that very few users are on monotherapy. Beyond that, there is also evidence that GLP-1RA have an endoplasmic stress-reducing effect ^51–53^. This may negate or overcome the diminished ER-stress response associated with reduced *XBP1* expression.

### Limitations

The use of BMI as a surrogate for body composition and high metabolic stress is a limitation. BMI has been shown to be less specific than measures such as waist-to-hip ratio, particularly in South Asians, who are more likely to carry fat ectopically ^54^. However, waist-to-hip ratio is not widely measured, and therefore not available across all study cohorts. Furthermore, BMI has been reliably shown to be associated with risk of diabetes and other cardiometabolic diseases across ethnicities ^39^ and given its wide availability, findings from this study can be used for risk stratification. Our use of ethnicity-specific BMI thresholds in part mitigates the misclassification of obesity. Furthermore, while we have undertaken robust replication, some well-known resources, such as the UK Biobank were not used as there under 2000 individuals of south Asian ancestry and 60 of East Asian ancestry with T2D before the application of any study inclusion criteria.

### Generalizability

Our findings are generalizable to the populations included in the study and have particular value to those with the phenotype of young onset diabetes in the absence of obesity. Comparing across ethnicities, individuals with young onset T2DM on average have higher HbA1c than later onset (**Supplementary Figure7**) ^1,2^. Of the sub-groups compared in this study, young + non-obese onset have worse glycaemic control at time of diagnosis compared to older + obese onset (**Supplementary Figure8**). People of South Asian descent are nearly twice as likely to have young onset T2DM and nearly four times more likely to have young onset with lean BMI compared to white Europeans ^3,6^.

A unique feature of this study is the use of ancestrally diverse populations, and in particular the dissection of South Asian ancestries. Consistent with population genetic studies of genetic diversity in South Asia ^55^, we observe differences across these two south Asian groups in risk allele frequency. In south Indians (DMDSC+TREND) we observe a higher percentage of homozygous risk allele carriers amongst compared to British Pakistani and Bangladeshis in G&H. The average age of diagnosis, BMI were lower in the South Indians, while HbA1c was higher **(Supplementary Table3).** Reflecting this diversity both in location and genetics, we have used different thresholds to classify non-obese BMI in non-migrant Asian Indians compared to migrant South Asians ^39,40^.

### Future directions

Our findings suggest that ethnicities such as South Asians have lower genetically-determined *XBP1* expression resulting in worse beta-cell response to metabolic stress leading to T2DM. Our findings support the value of genomics in precision diagnosis and therapeutics. Further studies of reversal of glycaemic deterioration for carriers of *XBP1* variant would be useful and could be undertaken using recall-by-genotype trials. The discovery of this novel association in type 2 diabetes and glycaemic traits in humans highlights the potential for development of strategies to therapeutically enhance *XBP1* expression in pancreatic beta cells to reverse beta-cell decline and dysfunction.

## Data Availability

All data produced in the present study are available upon reasonable request to the authors

## List of abbreviations

1000G: 1000 Genomes Project
CEU: Northern & Western European (ancestry group)
CHB: Han Chinese (ancestry group)
DIAMANTE: Diabetes Meta-Analysis of Trans-Ethnic association studies
DMDSC: Dr. Mohan’s Diabetes Specialties Centers
eQTL: expression Quantitative Trait Loci
ER: Endoplasmic Reticulum
G&H: Genes & Health
GWAS: Genome-Wide Association Study
HOMA-B: Homeostatic Model of Assessment - beta cell function
HOMA-S: Homeostatic Model of Assessment - insulin sensitivity
InsPIRE: Integrated network for systematic analysis of Pancreatic Islet RNA Expression
INSPIRED: INdia-Scotland PartnershIp for pREcision medicine in Diabetes
MAGIC: Meta-Analysis of Glucose and Insulin-related traits Consortium
T2DM: Type 2 Diabetes Mellitus
TDS: Tayside Diabetes Cohort, Scotland
TIGER: Translational human pancreatic Islet Genotype tissue-Expression Resource
TREND: Telemedicine Project for Screening diabetes and complications
UPR: Unfolded Protein Response
XBP1: X-Box Protein 1
YRI: Yoruba, Nigeria (ancestry group)
ZNRF3: Zinc and Ring Finger 3

## Acknowledgements

All volunteers who have facilitated this research across consortia and biobanks.

**Tayside Diabetes Cohort** (GoDARTS) has been funded and supported by the WTCCC (072960/Z/03/Z, 084726/Z/08/Z, 084727/Z/08/Z, 085475/Z/08/Z, 085475/B/08/Z) and as part of the European Union Innovative Medicines Initiative SUMMIT program. GoDARTS has been supported by Tenovus Scotland and Diabetes UK grants. SHARE is NHS Scotland Research infrastructure initiative and is funded by the Chief Scientist Office of the Scottish government. Additional Funding and initiation of the spare blood retention at NHS Tayside was supported by the Wellcome Trust Biomedical Resource Award (099177/Z/12/Z). Additional genome-wide array data were collected with funding from the National Institute for Health Research (INSPIRED [16/136/102]) with use of U.K. aid from the U.K. government to support global health research. We are grateful to all the participants in this study, the general practitioners, the Scottish School of Primary Care for their help in recruiting the participants and to the whole team, which includes interviewers, computer and laboratory technicians, clerical workers, research scientists, volunteers, managers, receptionists and nurses. The study complies with the Declaration of Helsinki. We acknowledge the support of the Health Informatics Centre, University of Dundee, for managing and supplying the anonymized data and NHS Tayside, the original data owner.

**TREND** and **DMDSC biobank** has been funded by the National Institute for Health Research (INSPIRED [16/136/102]) with use of U.K. aid from the U.K. government to support global health research

**Genes & Health** is/has recently been core-funded by Wellcome (WT102627, WT210561), the Medical Research Council (UK) (M009017, MR/X009777/1), Higher Education Funding Council for England Catalyst, Barts Charity (845/1796), Health Data Research UK (for London substantive site), and research delivery support from the NHS National Institute for Health Research Clinical Research Network (North Thames). Genes & Health is/has recently been funded by Alnylam Pharmaceuticals, Genomics PLC; and a Life Sciences Industry Consortium of Astra Zeneca PLC, Bristol-Myers Squibb Company, GlaxoSmithKline Research and Development Limited, Maze Therapeutics Inc, Merck Sharp & Dohme LLC, Novo Nordisk A/S, Pfizer Inc, Takeda Development Centre Americas Inc. We thank Social Action for Health, Centre of The Cell, members of our Community Advisory Group, and staff who have recruited and collected data from volunteers. We thank the NIHR National Biosample Centre (UK Biocentre), the Social Genetic & Developmental Psychiatry Centre (King’s College London), Wellcome Sanger Institute, and Broad Institute for sample processing, genotyping, sequencing and variant annotation. We thank: Barts Health NHS Trust, NHS Clinical Commissioning Groups (City and Hackney, Waltham Forest, Tower Hamlets, Newham, Redbridge, Havering, Barking and Dagenham), East London NHS Foundation Trust, Bradford Teaching Hospitals NHS Foundation Trust, Public Health England (especially David Wyllie), Discovery Data Service/Endeavour Health Charitable Trust (especially David Stables), NHS Digital - for GDPR-compliant data sharing backed by individual written informed consent. Most of all we thank all of the volunteers participating in **Genes & Health**.

For pharmacogenomic GWAS look-ups we would like to acknowledge the following individuals and consortia. For **Sulphonylurea response** Dr. Sook Wah Yee, Prof. Kathleen M Giacomini, and MetGen plus investigators. For **GLP1-RA response** IMI-DIRECT Investigators & GSK for the use of summary data from the HARMONY trials.

## Author contribution

TD: conceptualisation, formal analyses, visualisation, writing original draft and manuscript review. RMA: conceptualisation, supervision, data curation, project administration, resources, funding acquisition, manuscript review. SS, AYD, AM, MB, AT, ETA: data curation, formal analysis, methodology, visualisation, writing and reviewing manuscript. JS, VR, & RP: data curation, methodology, project administration, resources, manuscript revision. AS, DD, SH: data curation, formal analyses, validation, manuscript review. AM, JC, NS: methodology, manuscript review. RM, SF, AV: data curation, methodology, supervision, resources, validation, manuscript review. ERP, VM, CNAP: funding acquisition, project administration, methodology, resources, supervision, manuscript review. AAB: formal analysis, funding acquisition, investigation, methodology, supervision, visualisation, original draft, and review. MKS: data curation, formal analysis, funding acquisition, investigation, methodology, project administration, resources, supervision, visualisation, writing and review of manuscript.

## Ethical committee approvals

**DMDSC & TREND**: NIHR Global Health Research Unit on Global Diabetes Outcomes Research, Institutional Ethics Committee of Madras Diabetes Research Foundation, Chennai, India. IRB number IRB00002640. Granted 24^th^ August 2017.

**Tayside, Scotland UK**: Ethical approval for the study was provided by the Tayside Medical Ethics Committee (REF:053/04) and the study has been carried out in accordance with the Declaration of Helsinki.

**Genes & Health, UK**: Ethical approval is from the National Research Ethics Committee (London and Southeast), and the Health Research Authority (reference 13/LO/124), and Queen Mary University of London is Sponsor. All volunteers provide written informed consent, allowing analysis of health and genetic data and publication of results.

