## Supplementary Materials for "XBP1 expression in pancreatic islet cells is associated with poor glycaemic control across ancestries especially in young non-obese onset diabetes"

### Supplementary materials and methods

Supplementary Table 1. List of all data sources used, including information of data type, phenotype, and ancestry of study participants.

| **Data type** | **Data set name and acronym** | **Data level** | **Tissue/phenotype** | **Ancestry** |
| --- | --- | --- | --- | --- |
| Biobank: clinical and genetics | Tayside Diabetes Cohort (TDC)^1–3^ | Individual | HbA1c | White European (n=4908) |
| Biobank: clinical and genetics | Dr. Mohan’s Diabetes Specialty Centre (DMDSC)^4,5^ | Individual | HOMA-B, HbA1c | South Asian Indian (n=459) |
| Biobank: clinical and genetics | Telemedicine Project for Screening diabetes and complications  (TREND) | Individual | HbA1c | South Asian Indian (n=471) |
| Biobank: clinical and genetics | Genes & Health (East London Genes & Health)^6,7^ | Individual | HbA1c | South Asian Bangladeshi & Pakistani (n=644) |
| Transcriptomics | Gene-Tissue Expression (GTEx) consortium^8^ | Summary | Pancreas | White European  (n=305) |
| Transcriptomics | Integrated Network for Systematic analysis of Pancreatic Islet RNA Expression (InsPIRE) study^9^ | Summary | Pancreatic islets and beta cells | White European  (n_islets_=420, n_beta cells_=40) |
| Transcriptomics | Translational human pancreatic Islet Genotype tissue-Expression Resource TIGER) ^10^ | Summary | Pancreatic islet cells | White European  (n=399) |
| Genomics, GWAS | Diabetes Meta-Analysis of Trans-Ethnic association studies (DIAMANTE)^11^ | Summary | Type 2 diabetes | White European (n=251,739) |
| Genomics, GWAS | Biobank Japan ^12^ | Summary | Type 2 diabetes | East Asian Japanese (n=210,865) |
| Genomics, population selection | 1000 Genomes (1000G) selection browser ^13^ | Summary | Evidence of selection of gene region | White European, East Asian, West African |
| Genomics, GWAS | Meta-Analysis of Glucose and Insulin-related traits Consortium (MAGIC) ^14^ | Summary | HbA1c | White Europeans (n=132,400), East Asians (n=31,261) and South Asians (n=9525) |
| Pharmacogenomics, GWAS | MetGen Plus ^15^ | Summary | Sulphonylureas | White European  (n=5485) |
| Pharmacogenomics, GWAS | DIRECT ^16^ | Summary | GLP1-RA response | White European (predominantly)  (n=4571) |

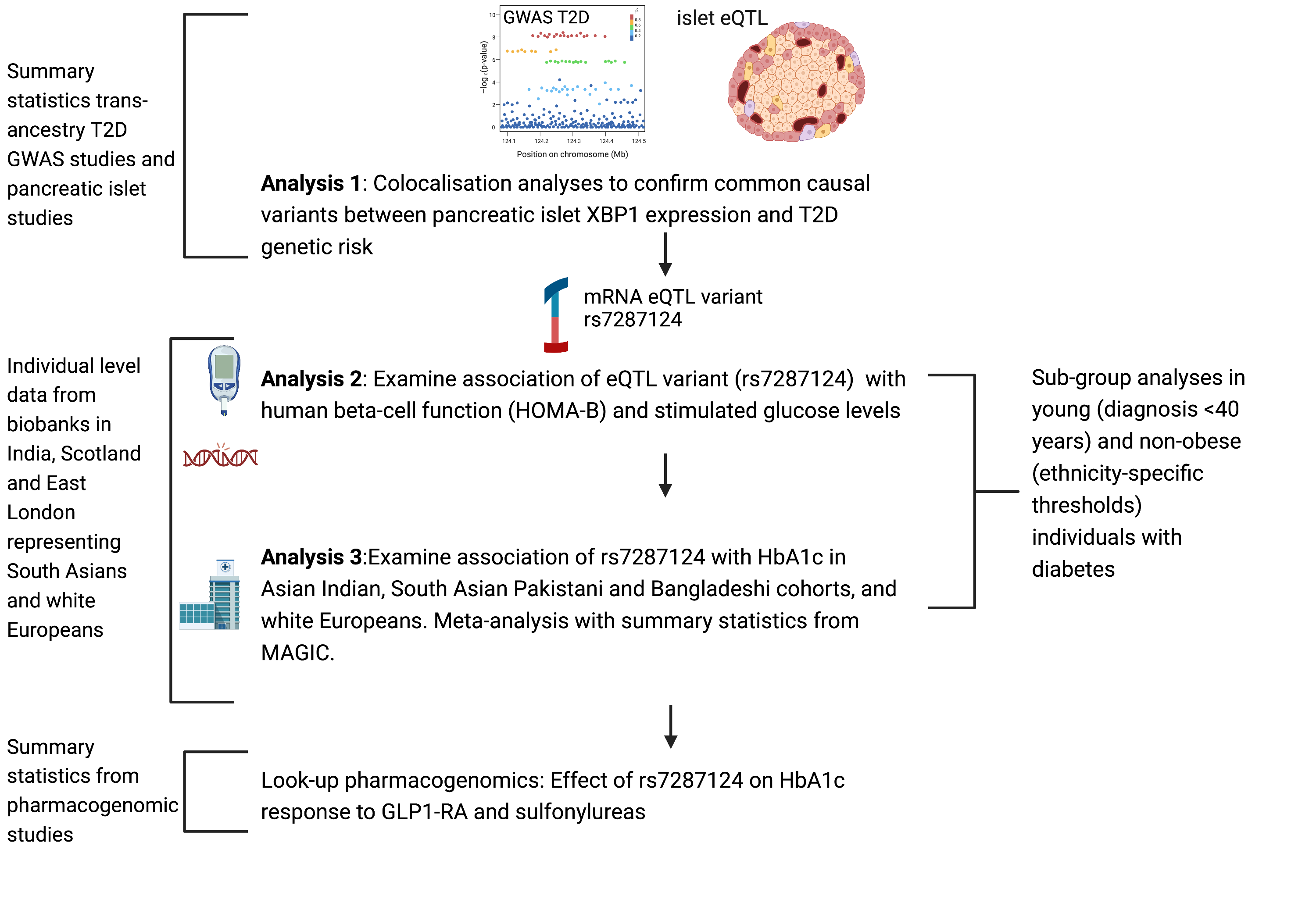

Supplementary Figure 1. Study flow-chart and analysis plan

#### Methods 1. Colocalisation analyses methods

##### Additional information of GTEx and InsPIRE for XBP1 expression

GTEx includes data from 305 pancreatic samples while InsPIRE included pancreatic islet samples from 420 donors ^8,9^. We used summary statistics (Z score) to identify the variants with the largest and most significant effects on XBP1 expression from these two resources. In InsPIRE, we used expresion data from all islet cells, and having identified a putative lead eQTL, confirmed the direction of effect in beta-cells. Then using the TIGER resource (Translational human pancreatic Islet Genotype tissue-Expression Resource TIGER) we confirm the same variant was a lead eQTL for XBP1 in islets and that it did not have any *cis* or *trans* effects on expression of any other genes (also see Supplementary Result 1, Table 1).

##### Genome-wide association summary statistics of type 2 diabetes

We used summary statistics from two well-powered type 2 diabetes GWAS, one containing individuals of white European ancestry from Diabetes Meta-Analysis of Trans-Ethnic association studies (DIAMANTE) and the second from those of East Asian ancestry from Biobank Japan ^17^. We used LD structures of the respective GWAS populations and a standard 10 kb window around the transcription start site of *XBP1*. DIAMANTE included over 180,000 T2DM cases and 1.1 million controls with 48.9% being of non-European descent.

#### Methods 2. Study biobank details

###### Table 1. INSPIRED – India-Scotland Partnership for Precision Medicine in Diabetes

The National Institutes of Health Research (NIHR)-funded the INSPIRED project to better understand the differences in genetics of type 2 diabetes across these two ethnicities. Through this a biobank of 26,000 genotyped Asian Indians and 15,000 white Scottish individuals was created. The specific INSPIRED cohorts are detailed below. Genotyping was carried out using the Illumina Global Screening Array for the two cohorts from India, G&H, and a large proportion of TDC.

1. Dr. Mohan’s Diabetes Specialities Centre (INSPIRED-DMDSC)

DDMSC is a privately-run chain of specialty hospitals across India. DMDSC has a single electronic medical record-keeping system to record patient encounters, biochemistry testing, retinal screening etc. These records were linked to genotype data created through NIHR-INSPIRED for the study population. Any individuals who were diagnosed with type 1 diabetes at diagnosis or at any point during follow-up care or who tested positive for GAD65 antibodies were excluded for analysis. Health records were queried for HbA1c tests and BMI recorded during the study period (1 year before and up to 1 months after diagnosis of T2D).

###### 2. Telemedicine Project for Screening diabetes and complications in rural Tamilnadu (INSPIRED-TREND)

This Telemedicine PRoject for screENing Diabetes and its complications in rural Tamil Nadu (TREND) cohort (n=11,293), conducted in 30 villages of Cheyur Taluk in Chengalpattu/Kancheepuram districts of Tamil Nadu, India between 2018 and 2021. Of the total population screened and genotyped, 1,637 had T2D (of which 1154 had known diabetes and 480 were diagnosed during the survey). During the survey anthropometric data: height, weight, bioimpedance based body fat measurements, oral glucose tolerance tests and HbA1c (HbA1c only in those with OGTT indicating T2D). Additional details on the survey methods are described elsewhere ^18^. In all participants, an oral glucose tolerance test (OGTT) using a venous blood sample was done by administering 75g of anhydrous glucose dissolved in 200 ml of water, after 10-12 hours of overnight fasting (except in individuals with self-reported diabetes, for whom only fasting plasma glucose was done). A fasting venous plasma glucose sample and 2-hour post-load glucose samples were collected. Individuals surveyed and found to have newly diagnosed diabetes were included in this analysis; HbA1c, BMI and age recorded at the time of the survey were considered to be features at diagnosis.

INSPIRED-DMDSC and TREND genotyping was performed using Illumina Global Screening Array (GSA) and imputation was performed using the Haplotype Reference Consortium ^19^

###### 3. Tayside Diabetes Cohort – East of Scotland Diabetes Cohort (TDC)

TDS is based at the University of Dundee, UK. Combining the GoDARTS and SHARE cohorts which both have electronic health record linkage through NHS Tayside and Fife ^2,3^. Linkage of health data particularly for diabetes diagnosis and management in this resource is well-documented ^1^. Historic genotyping before 2015 (n=8000 T2D) was performed using Affymetrix 6.0 and Illiumina HumanOmni Express. Recent genotyping for this resource (n-5000 T2D) has been undertaken using the Illumina GSA panel. All imputation was performed using HRC. Using Scottish Diabetes Care Network the date of diagnosis is recorded and can be triangulated with BMI assessments and HbA1c measurements using clinical and biochemistry records respectively.

###### 4. East London Genes & Health (G&H)

G&H are based in Queen Mary University of London, UK and is one the largest resources globally for the study of migrant British Pakistani and British Bangladeshi populations ^6^. Linkage to electronic medical records from local NHS boards enables us to determine the date of diabetes diagnosis and find the closest measures of BMI and HbA1c per study design.

##### Genetic data

Genotyping data was available for all biobanks and genotyping methods of each have been described previously ^1,2,4,7^. Genotyping for G&H, DMDSC, and TREND was performed using the Illumina Global Screening Array (GSA) chip, whereas TDS used a combination of GSA and historic Affymetrix Illumina genome-wide arrays. Imputation was performed using the Michigan Imputation Server for all biobanks. G&H imputation was performed using the Genome Asia panel ^16^, whereas imputation for TDS, DMDSC and TREND were undertaken using the Haplotype Reference Consortium panel ^9^. Quality control measures for these data have also been described previously ^7,20^, briefly the quality of imputation for the variant was high across all data sources and any cases with imputation quality < 95% were discarded. Relatedness tests were undertaken in all cohorts and all first-degree relatives were excluded.

#### Supplementary Results

#### Result 1. eQTL variant for XBP1 expression in pancreatic islet cells

Supplementary Table 2. TIGER output showing pancreatic islet eQTL associations with this variant suggesting it is only an eQTL for XBP1 expression

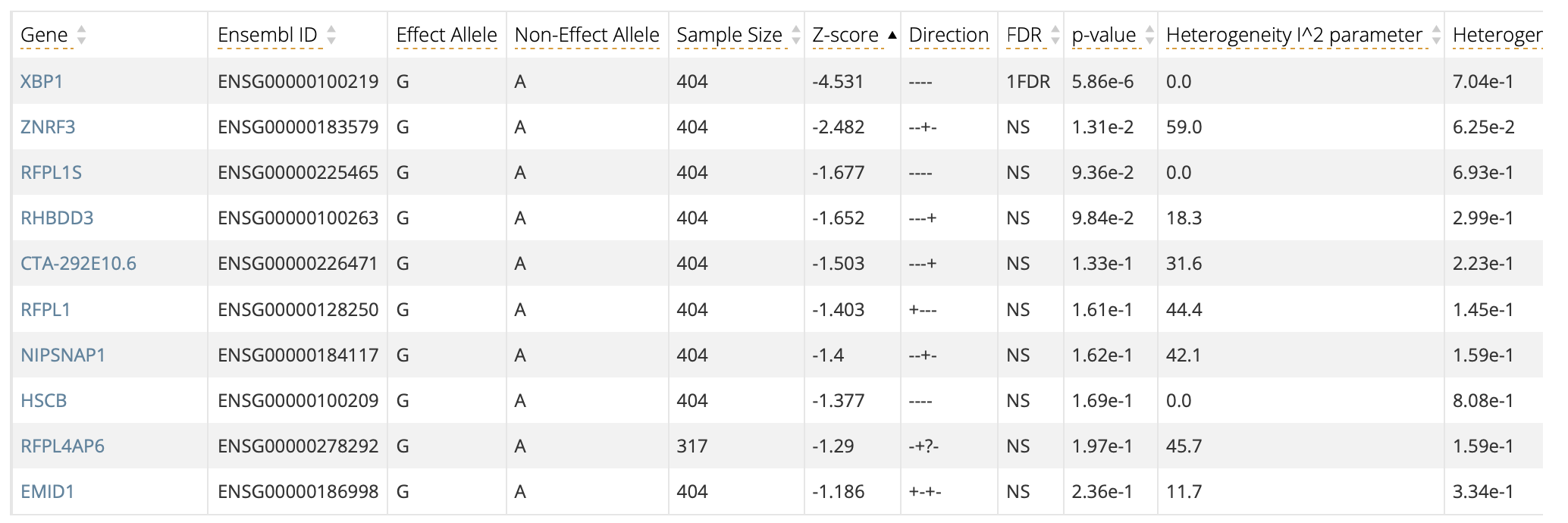

#### Result 2. Colocalisation analyses

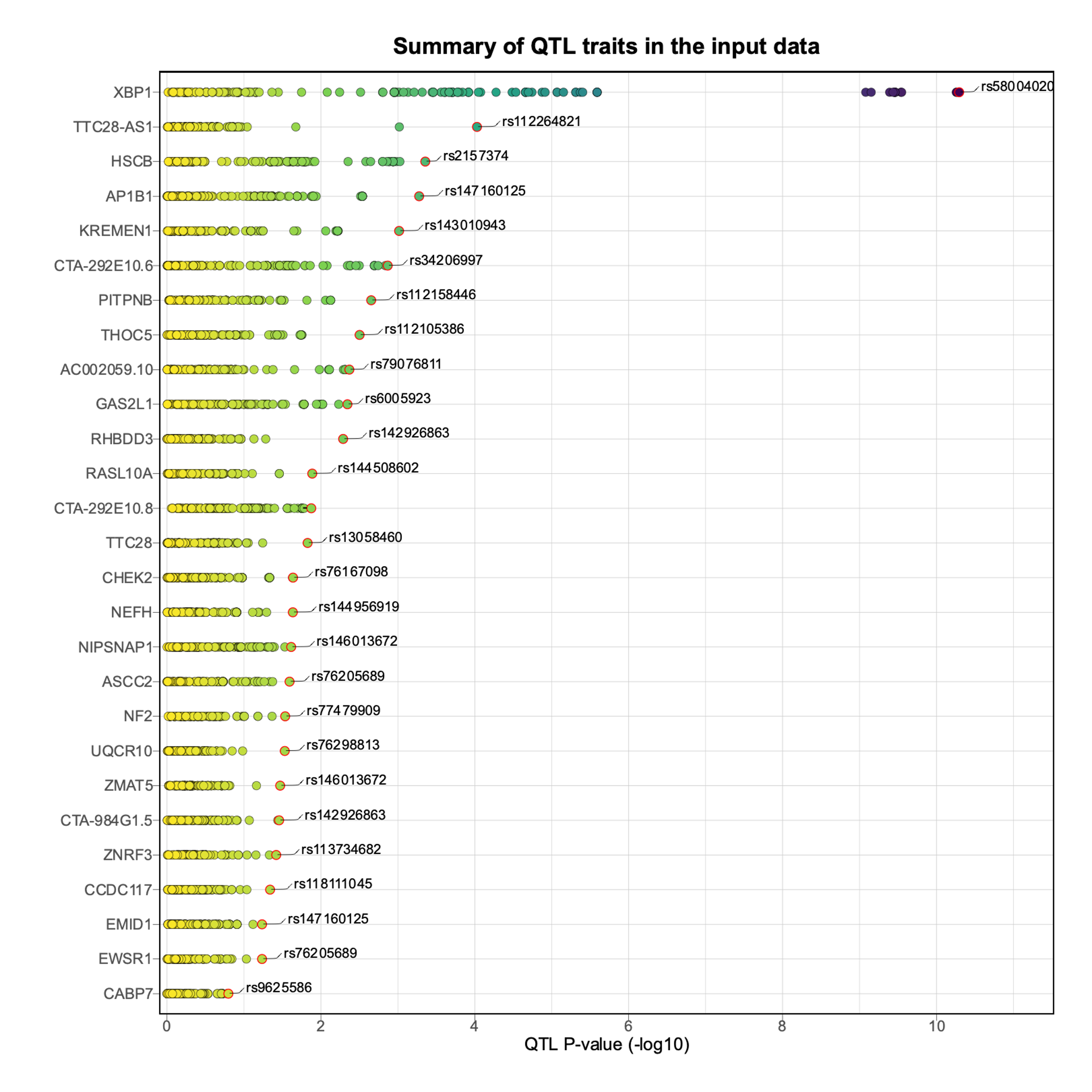

Supplementary Figure 2. The plot produced by ezQTL above shows the summary of association P-value of each trait in the QTL dataset. The QTL traits are sorted according to the most significant P-value. The top significant associated variants for each trait are highlighted and labelled with rsID. The color-coding of the variants was based on P-value scale. Due to figure size limitation, only the top 50 traits are included in this figure.

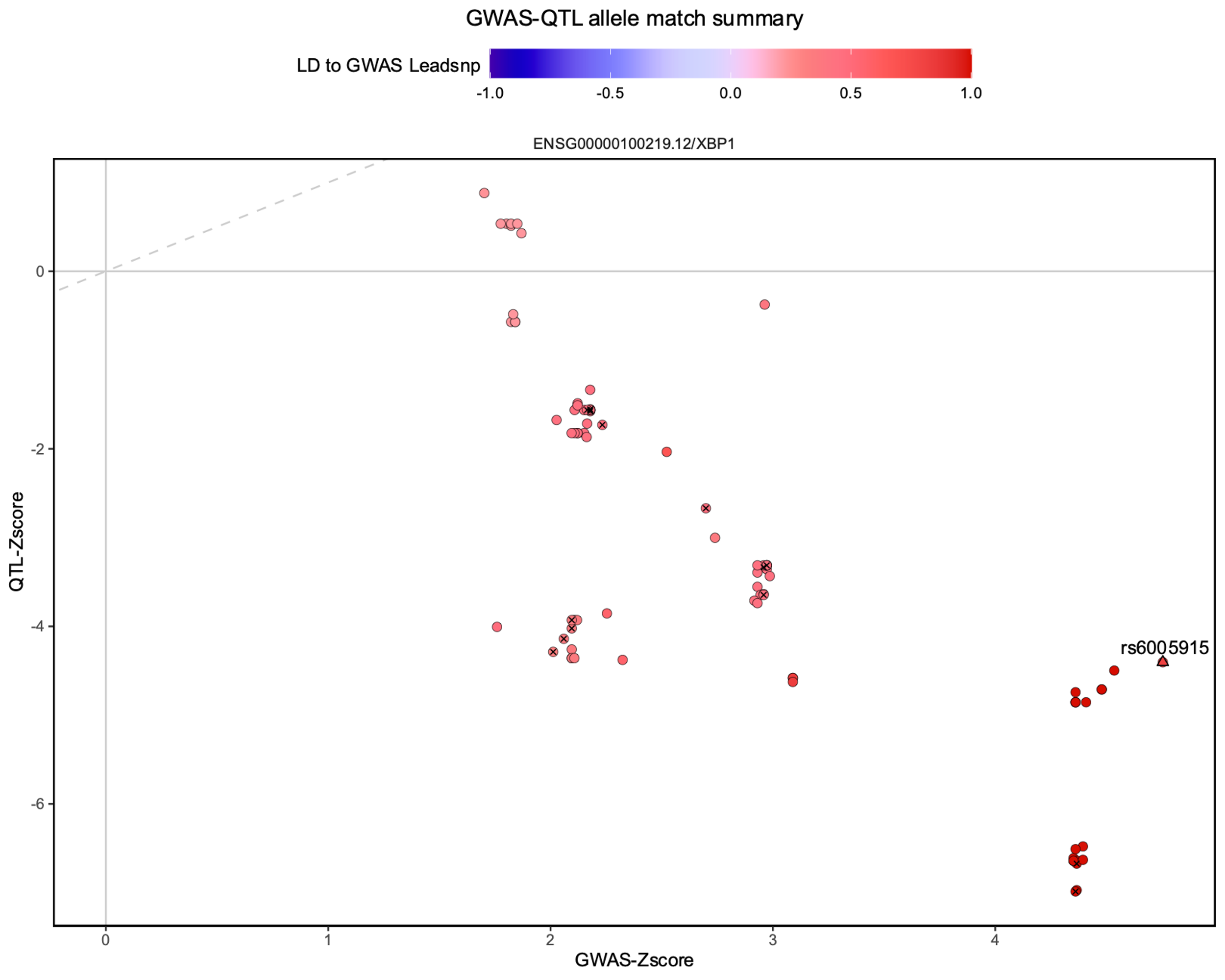

Supplementary Figure 3. Scatterplot shows the correlation of Z-scores between QTL and GWAS data. For the matched alleles in both the QTL and GWAS datasets, a strong positive or negative correlation is expected for a colocalized locus.

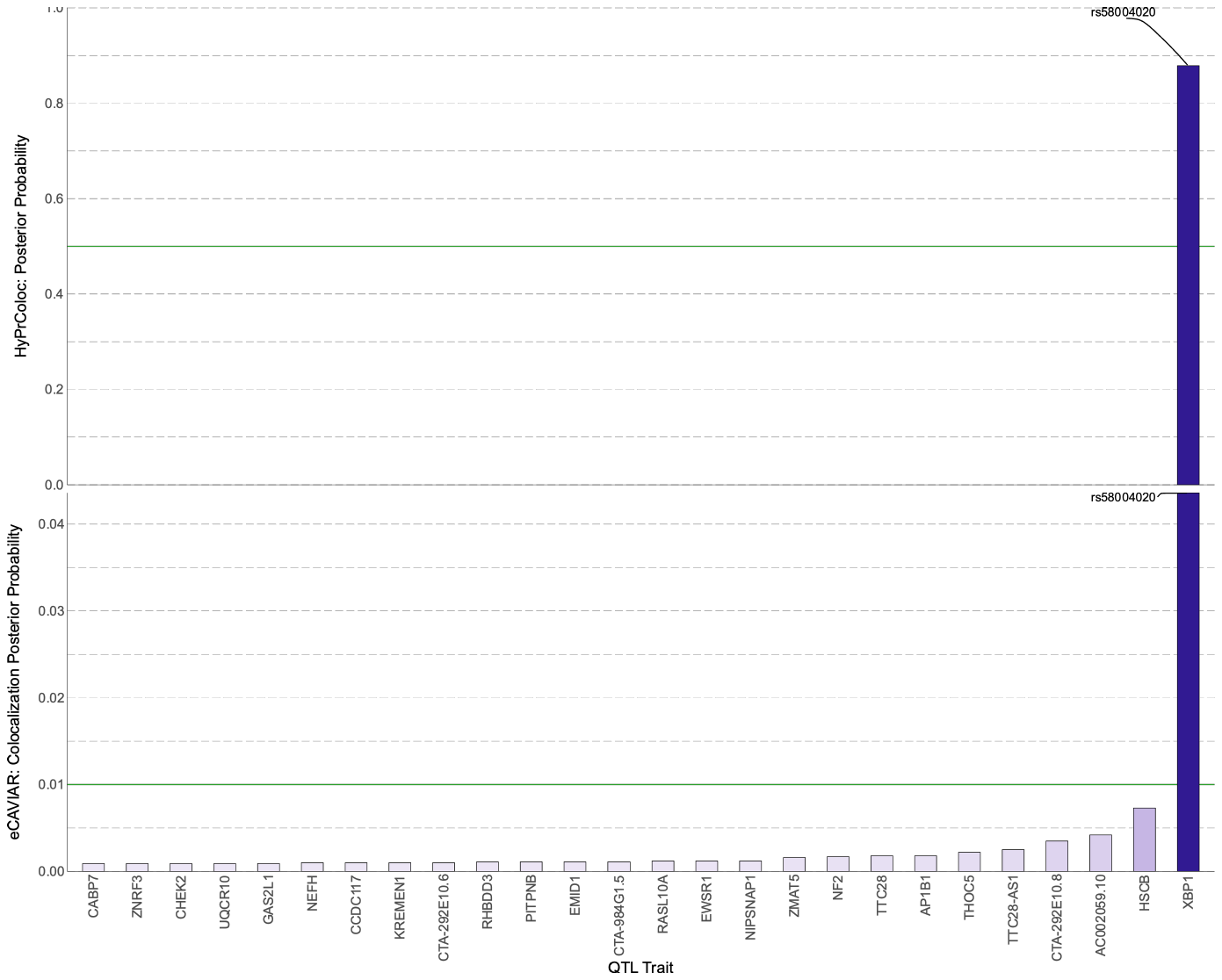

Supplementary Figure 4. The colocalisation analyses of mRNA expression using HyPrColoc (upper panel) and eCAVIAR (lower panel) consistently show that XBP1 expression in the pancreatic islets and type 2 diabetes risk are colocalised in East Asians. Any trait that crosses the solid green line, representing the threshold recommended by the specific method, is considered significant. Both methods find XBP1 to be a causal gene. A potential causal variant for expression is identified, rs58004020, which is located gene ZNRF3 and is a cis eQTL for XBP1. In East Asians, R2=0.99, D’=1 between rs58004020 and islet eQTL rs7287124.

Supplementary Table 3. Results of colocalisation analyses of XBP1 expression and type 2 diabetes risk

|  | | | Ethnicity | | | |
| --- | --- | --- | --- | --- | --- | --- |
|  |  |  | East Asians | | White Europeans | |
|  |  |  | CLPP | PP | CLPP | PP |
| eQTL source |  | Method |  |  |  |  |
|  | InsPIRE  Pancreatic islets | HyPrColoc | NA | 0.88 | NA | 0 |
|  |  | eCAVIAR | 0.01 | NA | 0.003 | NA |
|  | GTEx pancreas | HyPrColoc | NA | 0.87 | NA | 0 |
|  |  | eCAVIAR | 0.05 | NA | 0.0055 | NA |

Colocalisation analyses were performed using data from the InsPIRE pancreatic islets, GTEx consortoium pancreatic tissue samples and type 2 diabetes (T2D) GWAS from Biobank Japan and DIAMANTE (white Europeans). High probability of colocalisation for expression and T2D risk was observed from XBP1 expression in the pancreas for East Asians from Biobank Japan, with more modest effects noted in for expression in islets. Both analyses identified variants in the gene ZNRF3, which neighbours XBP1 and contains eQTLs for XBP1.

#### Human Genetic Evidence (HuGE) calculator results for *ZNRF3* and *TCF7L2*

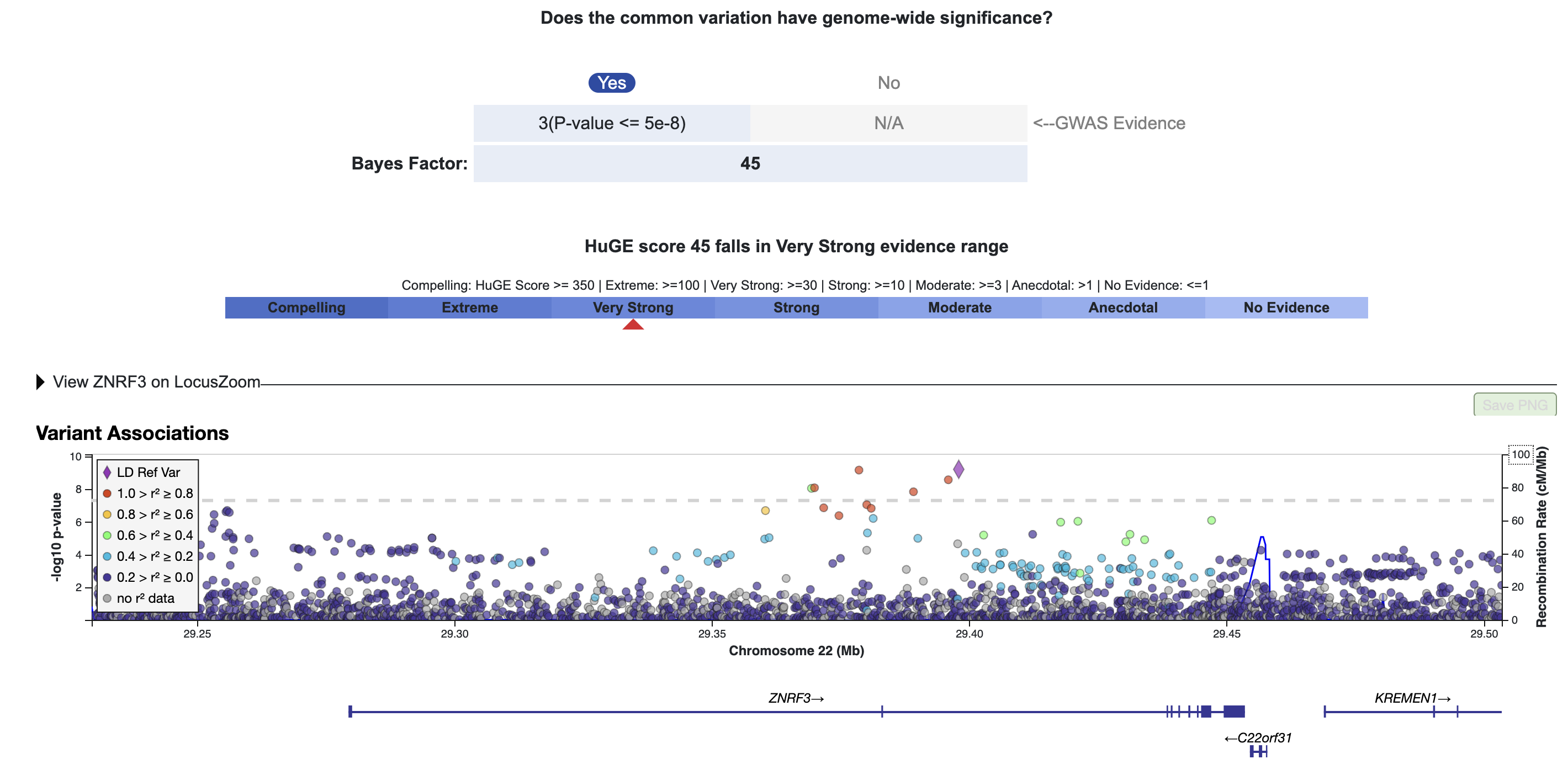

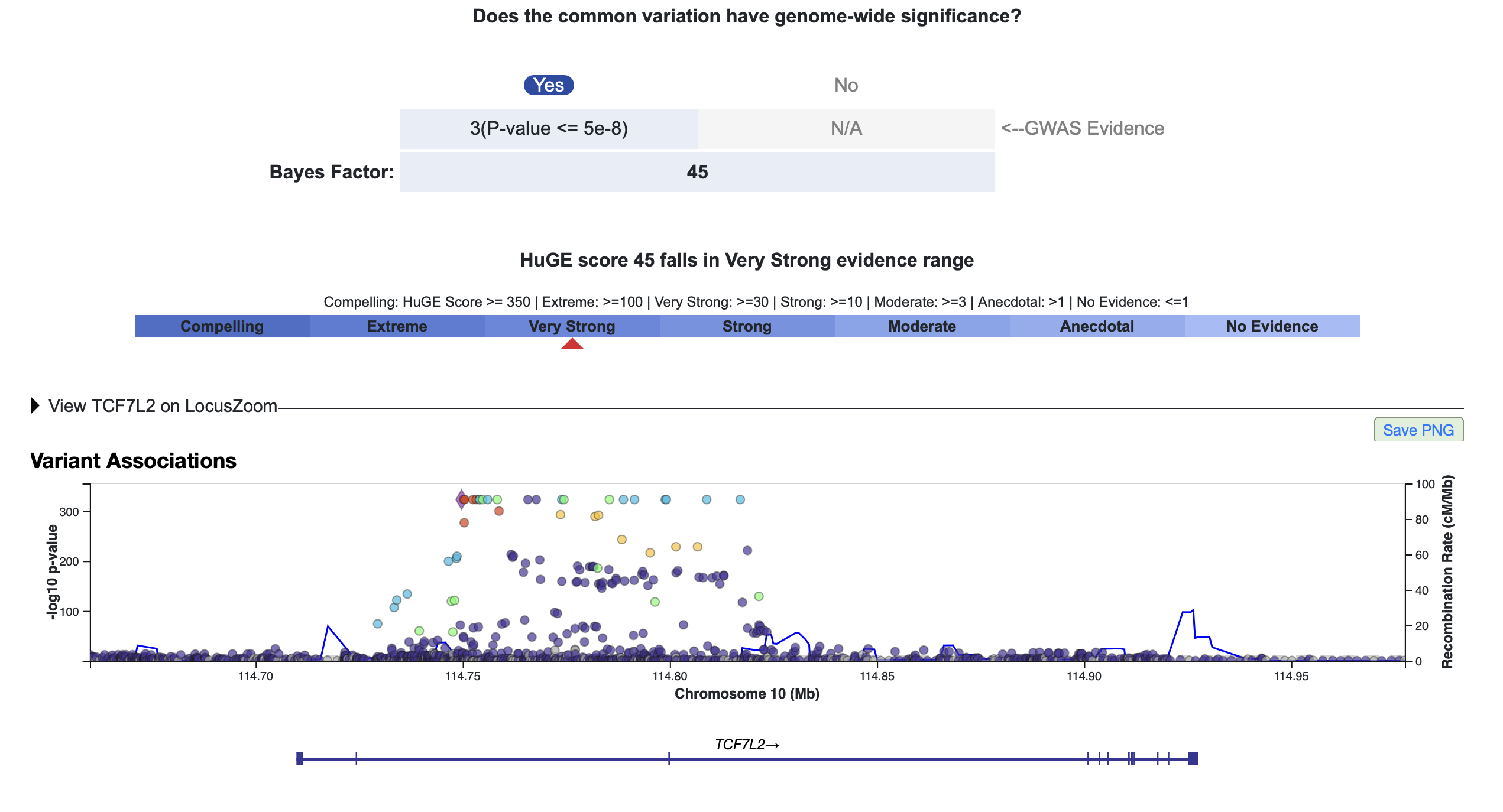

Supplementary Figure 5. Results of HuGE analysis for evidence of genetic support from ZNRF3 (contains eQTLs for XBP1) (upper panel) and TCF7L2 (lower panel) with type 2 diabetes. The HuGE score suggests a very strong level of evidence that common variants ZNRF3 are associated with type 2 diabetes risk. There was no evidence of involvement of rare variants in the gene. The Bayes Factor for the well-known T2D gene TCF7L2 also suggested the same level of evidence for type 2 diabetes.

#### Result 2. Cohort information and analyses

Supplementary Table 4. Descriptive statistics of the cohorts used

| Ethnicity | Asian Indian | | White European | South Asian (British Pakistani and Bangladeshi) |
| --- | --- | --- | --- | --- |
|  | INSPIRED-TREND | INSPIRED-DMDSC | INSPIRED-TDS | Genes & Health |
| N, mean and SD | 471 | 459 | 4908 | 644 |
| Age at Diagnosis (years) | 48.9 (12.4) | 44.2 (11.6) | 61.5 (11.9) | 47.7 (10.9) |
| BMI (kg/m^2^) | 25.7 (4.6) | 26.7 (4.4) | 32.0 (6.6) | 28.8 (5.0) |
| HbA1c (%) | 8.8 (2.5) | 9.2 (2.4) | 8.5 (2.3) | 7.0 (1.5) |
| HbA1c (mmol/mol) | 69.5 (25.6) | 77.0 (25.9) | 69.3 (25.1) | 53.14 (16.2) |
| Sex (% Female) | 47% | 40% | 45% | 46% |
| EAF rs7287124 | 0.50 | 0.49 | 0.22 | 0.46 |
| EAF rs7287124 Young & non-obese type 2 diabetes | 0.52 | 0.52 | 0.26 | 0.48 |

INSPIRED: India Scotland Partnership for Precision Medicine in Diabetes; TREND: Telemedicine Project for Screening diabetes and complications in rural Tamilnadu; DMDSC: Dr. Mohan’s Diabetes Specialties Clinic; TDS: Tayside Diabetes Study; Genes & Health: East London Genes and Health Study. EAF: Effect Allele Frequency

Supplementary Table 5. Association between log-transformed HOMA-B and XBP1 eQTL variant rs7287124 in additive and recessive models. These two genetic models were tested based on Figure 3 which shows a slightly recessive effect in unadjusted models

|  | N | B _recessive_, SE_recessive_ | P _recessive_ | B _additive_,  SE _additive_ | P _additive_ |
| --- | --- | --- | --- | --- | --- |
| Main effects models | | | | | |
| HOMA-B overall | 470 | -0.26, 0.09 | 0.003 | -0.11, 0.054 | 0.033 |
| HOMA-B in young and non-obese | 82 | -0.60, 0.24 | 0.014 | -0.32, 0.14 | 0.024 |
| HOMA-B in older and obese | 134 | -0.16, 0.17 | 0.34 | -0.10, 0.12 | 0.30 |
| Adjusted for HOMAS, age, and sex | | | | | |
| HOMA-B overall | 470 | -0.267, 0.087 | 0.0025 | -0.14, 0.05 | 0.005 |
| HOMA-B in young and non-obese | 82 | -0.56, 0.24 | 0.02 | -0.30, 0.14 | 0.036 |
| HOMA-B in older and obese | 134 | -0.18, 0.17 | 0.27 | -0.11, 0.10 | 0.27 |

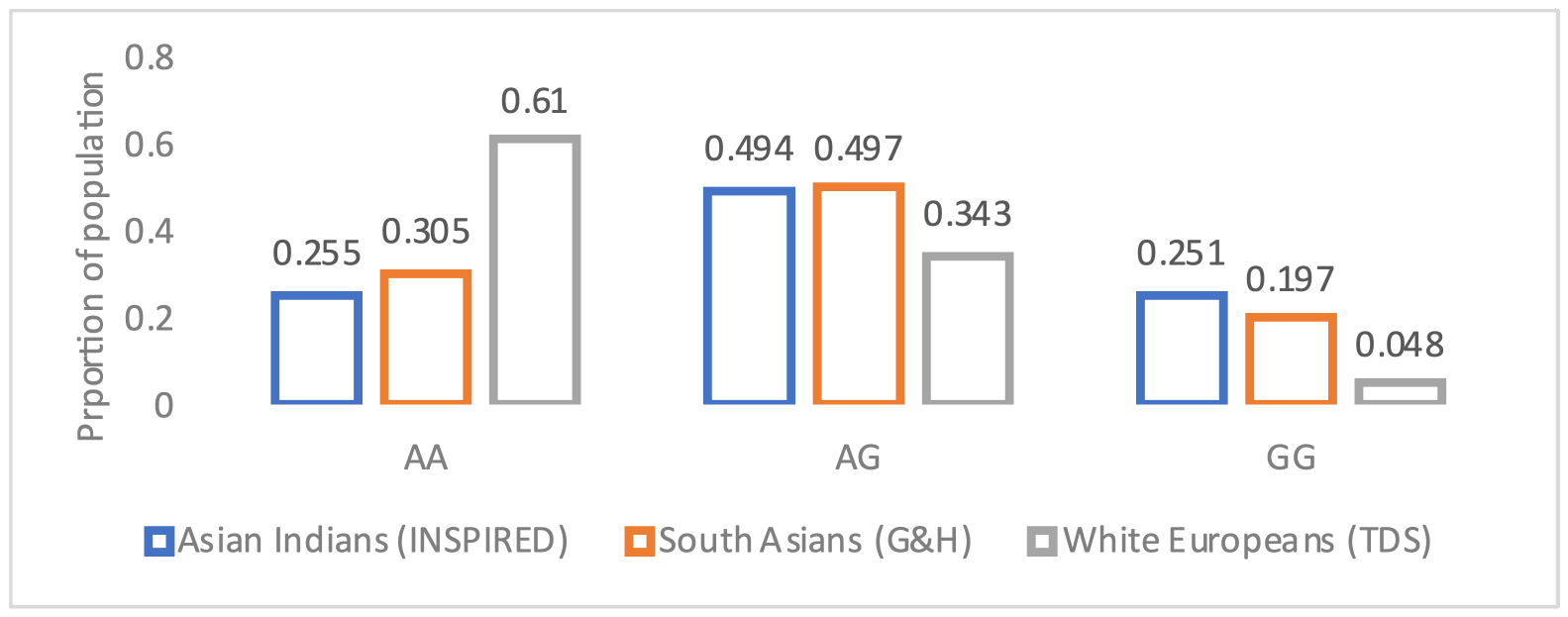

Supplementary Figure 6. The proportion of the population in study cohorts carrying each genotype of rs7287124 across 3 study sites. The proportion carrying the GG alleles are at greatest risk of poor glycaemic control. While the MAF in Asian Indians and South Asians is between 50% an 46% respectively, the underlying frequency of homozygosity is 25% and 20% respectively, compared to 0.05% in white Europeans.

#### Result 5. Pharmacogenetic effects of *XBP1* eQTL variant rs7287124 on sulphonylurea and GLP1-RA response

Supplementary Table 6. Association of rs7287124 with HbA1c response to sulphonylureas and GLP-1RA

| Sulphonylurea response (n=5,485) ^15^ (MetGen consortium) | | | | |
| --- | --- | --- | --- | --- |
| Variant | EA/NEA | EAF | Beta ± SE (mmol/mol) | P |
| rs7287124 | G/A | 0.247 | -0.061 ± 0.023 | 0.008 |
| GLP-1RA (n=4,463) ^16^ (DIRECT consortium) | | | | |
| rs7287124 | G/A | 0.28 | 0.019, 0.022 | 0.42 |

MetGen: EA: Effect allele, NEA: Non-effect allele, SE: standard error, P: P value, HbA1c effects provided in % as per original publications.

#### Discussion

#### Section 6. Population selection

#### Method: genetic selection of *XBP1* in human populations

To determine population selection, a score for evidence of recent positive selection was calculated for 26,506 protein coding and linc RNA genes, based on the gencode 26 annotation 6. This score was based on the Cross Population Extended Haplotype Homozygosity (XPEHH) 7 score looking for evidence of greater recent positive selection in a population of European ancestry (the CEU population from 1000 genomes) relative to one of East Asian (CHB) or Yoruban ancestry (YRI). A gene score was calculated by averaging the selection score at every position in a window 20kb up and downstream of the transcription start site of the gene with P<0.05 or taking the maximum score if no positions were significant. This window contains both *XBP1* and *ZNRF3*. These scores were ranked to produce a list of genes with the greatest evidence that the gene had been under greater recent selection pressure in each pairwise combination of populations.

#### Result*: XBP1* under positive selection in Europeans

The observed allele frequencies are a function of population history, and the lower frequencies of risk alleles we observe in European populations could be the result of protective alleles being positively selected for in individuals of European ancestry. Indeed, calculating a gene-based score for recent selection based on data from the 1000 Genomes Selection Browser ^37^, we find *XBP1* (and a 20kb region flanking it which includes *ZNRF3*) is ranked in the top 6.8% of all genes and long non-coding RNAs in terms of evidence of recent selection in the European CEU population relative to the Chinese CHB population **(Supplementary Methods 2**). There is weaker evidence of selection in the CEU population relative to the Yoruba population in Nigeria (ranked in the top 28.1%) but no evidence of differing selection pressures between YRI and CHB (gene ranked 94.3%). Genetic selection across ancestries helps identify genes with potentially important functional roles, with recent selection suggesting an advantage to preserving biological function ^50^. These results are consistent with our hypothesis that protective alleles have been selected for to a greater degree in European populations compared to East Asian populations.

#### Discussion: BMI and HbA1c in younger v. older onset type 2 diabetes in INSPIRED cohorts

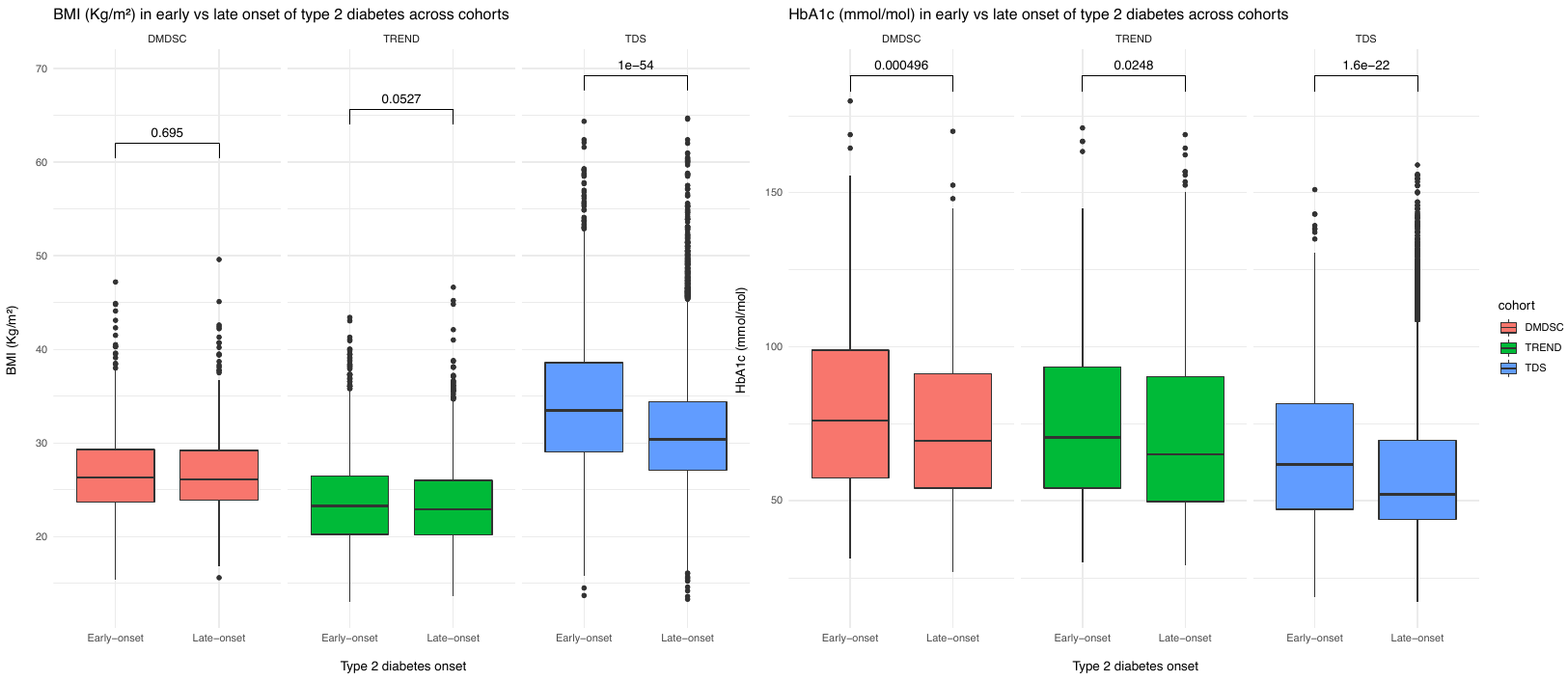

Supplementary Figure 7. HbA1c in those with early v. later onset T2D across cohorts. HbA1c is consistently higher in those with early onset T2D.

#### Discussion: HbA1c in young and non-obese onset type 2 diabetes in INSPIRED cohorts

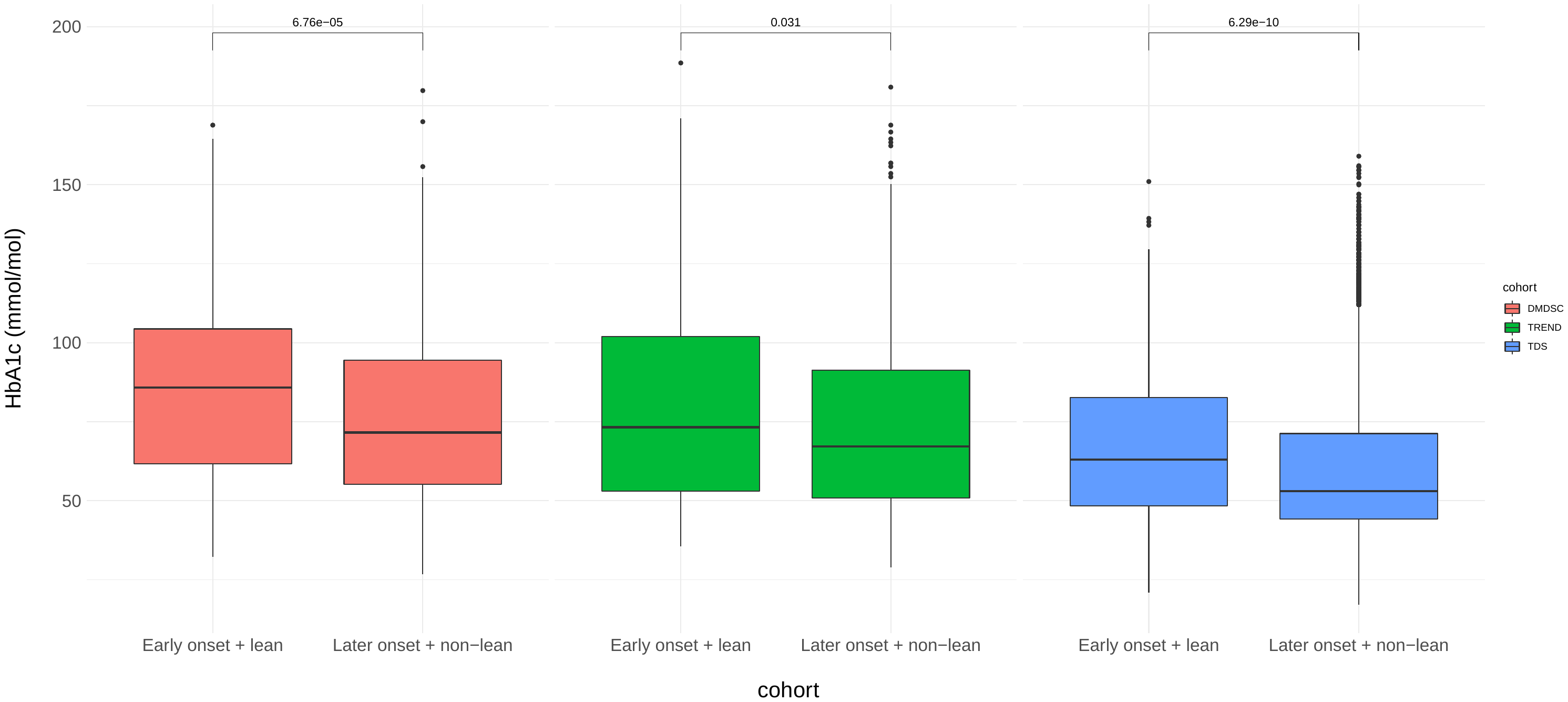

Supplementary Figure 8. Native or pre-treatment HbA1c levels in those diagnosed young and non-obese compared to later onset with obese BMI across the cohorts. Individuals with young and non-obese onset consistently have significantly higher HbA1c.
